# Location of joint involvement differentiates rheumatoid arthritis into different clinical subsets

**DOI:** 10.1101/2023.09.19.23295482

**Authors:** Tjardo D. Maarseveen, Marc P. Maurits, Stefan Böhringer, Nils Steinz, Sytske Anne Bergstra, Bianca Boxma-de Klerk, Herman Kasper Glas, Josien Veris-van Dieren, Annette H. M. van der Helm-van Mil, Cornelia F. Allaart, Marcel J T. Reinders, Tom W J Huizinga, Erik B van den Akker, Rachel Knevel

## Abstract

**Objectives:** To aid research on etiology and treatment of the heterogeneous rheumatoid arthritis (RA) population, we aimed to identify phenotypically distinct RA subsets using baseline clinical data.

**Method:** We collected hematology, serology, joint location, age and sex of RA-patients from the Leiden Rheumatology clinic(n=1,387). We used deep learning and clustering to identify phenotypically distinct RA subsets. To ensure robustness, we tested a) cluster stability, b) physician impact, c) association with remission and methotrexate failure, d) replication in clinical trial data (n=307) and independent secondary care (9 clinics, n=515).

**Results:** We identified four subsets: Cluster-1) arthritis in feet, Cluster-2) seropositive oligo-articular disease, Cluster-3) seronegative hand arthritis, and Cluster-4) polyarthritis. We found high cluster stability, no physician influence, significant difference in methotrexate failure(*P*<0.001) and occurrence of remission*(P*=0.007). The MTX-failure rates were recurrent in both replication sets(both *P*<0.001).

The hand-Cluster-3 showed best outcomes, especially compared to Cluster-4 (*P*<0.001) and Cluster-1 (*P*=0.003). The MTX-failure difference was largest in the ACPA-positive stratum (Cluster-3 versus Cluster-1 (HR:0.3 (0.15-0.60) *P*<0.001), Cluster-3 versus Cluster-4 HR:0.33 (0.15-0.72) *P*=0.005). We observed this for Cluster-4 in all sets, and for Cluster-1 in two out of three. This was independent of baseline disease activity and symptom duration. The clusters significantly improved the MTX failure model on top of traditional risk factors.

**Conclusions:** We discovered and replicated four phenotypic subgroups of RA at baseline characterized by hand and foot involvement that associate with treatment response. Such knowledge on disease subgroups could enhance studies to the treatment and the mechanisms underlying RA.

**KEY MESSAGES:** *What is already known about the subject?*

- Rheumatoid arthritis is a heterogeneous disease and clinicians have not completely identified the disease differentiating patterns in clinical practice.
- Data-driven unsupervised techniques are able to identify hidden structures in big data.

*What does this study add?*

- We identified four RA clusters at baseline: feet involvement, oligo-articular disease, hand involvement and polyarthritis (both feet and hand involvement). The hand cluster shows a good treatment response, especially when compared to the polyarthritis and feet group.
- The difference in treatment success between hands vs foot clusters was largest in the ACPA-positive stratum.

*How might this impact on clinical practice or future developments?*.

- This research supports future endeavors in identifying etiological mechanisms and tailored treatment solutions.
- Feet involvement at disease presentation can be considered a risk factor for less successful treatment.
- Depending on the location of inflamed joints at first presentation, RA-patients might need different treatment.

## INTRODUCTION

Rheumatoid arthritis (RA) is a heterogeneous disease. The current classification criteria for RA were developed to approximate the decision to start early treatment and the exclusion of other diseases. At clinical presentation, patients vary in the number and pattern of joints involved, presence of extra articular manifestations and abnormalities in blood and synovial fluid [1, 2]. The heterogeneity of RA also manifests in clinical outcomes, namely prognosis, treatment response and comorbidities. This evident diversity likely impacts the interpretation of treatment effect and etiologic factors such as genetics and downplay their importance altogether [3]. If phenotypic subsetting into more homogeneous groups is possible, there is a potential for better research into the etiology and enhancing the treatment of RA.

For centuries, pattern recognition on clinical variables by doctors has been the driving force of disease identification and examination of the underlying etiologic mechanisms. Thus far clinicians have not identified the relevant (sub)patterns in RA. The presence of ACPA [4, 5, 6, 7] and the age of onset [8, 9, 10] have been raised as possible dichotomous disease subsetting features. However, neither of these markers in isolation adequately addresses the heterogeneity and complexity of the disease. This suggests there are other factors involved.

Cluster analysis combining a high number of factors has demonstrated its effectiveness in categorizing complex diseases (such as diabetes type II, asthma, osteoarthritis) into subtypes that differ in clinical outcomes or biological background [11, 12, 13]. In the context of RA, there is quite some focus on molecular phenotyping such as done by Lewis et al [14], who discovered patterns in synovial tissue at baseline, with the lymphoid-myeloid pathotype being a predictor for a poor outcome at disease onset [15]. Others used clinical and comorbidity information for clustering and identified four subsets, including one that exhibited a higher likelihood for biological DMARD initiation [16]. Likewise, Curtis et al [17] used clinical variables, though not exclusively at baseline, and identified five clusters that differed in disease activity, RA-duration and type of comorbidities. These outcomes are typically highly influenced by treatment decisions and events that occur independent of the specific RA type. Furthermore, detailed clinical information such as the pattern of involved joints may be relevant for disease differentiation as exemplified by psoriatic arthritis [18], yet none of the previous studies capitalize on this information for clustering.

Electronic Health Records (EHR) data provides a powerful asset for clustering as it encompasses a wide variety of data types (laboratory values, clinical examination, demographics) that each offer a unique perspective on the patient’s condition. The EHRs are collected as routine clinical care, and thus resemble the true patient population more closely than a study population collected with a particular hypothesis in mind. The diversity of data types does however pose a methodological challenge due to structural differences between the data modalities. The recent surge of deep learning tools [19], offers the possibility to combine different EHR-layers into a patient representation by extracting the (hidden) factors that capture most variation in the data. At present, there exist many machine learning (ML) techniques to learn the relevant (clinical) patterns, and encode patients accordingly. These embeddings can be used to detect patients’ subgroups, identify patterns, build predictive models or assist in making disease classifications. The literature reports that clustering on top of these embeddings typically outperforms conventional techniques in the case of high dimensional or complicated data [20, 21, 22].

In this study we aimed to illuminate the clinical heterogeneity within RA by using the symptoms at initial presentation so before external factors such as treatment interfere. We hypothesize that the location of the involved joints and the inflammatory patterns observed in the blood play a role in subsetting RA, similar to their significance in distinguishing PsA from RA [18]. To achieve this we make use of state-of-the-art data-driven techniques to identify the disease-differentiating signatures of RA using baseline clinical variables.

## METHODS

### Patients

Our study comprises two different phases (Fig. S1): a developmental phase to identify the subtypes at baseline in a training set (set A) and a validation phase that uses historic trial data (set B) and external data from regional centers (set C).

Set A consisted of RA patients that visited the rheumatology outpatient clinic of the Leiden University Medical Centre (LUMC) for the first time between August 29th, 2011 till December 1st 2022. RA diagnosis was based on the physician’s diagnosis within 1 year since first visit.[23, 24].

Set B concerned RA patients from the IMPROVED trial that were recruited between March 2007 till September 2010 [25]. This trial recruited undifferentiated arthritis and early RA with less than 2 years of symptoms. We selected only those patients who met the ACR2010 criteria within one year after inclusion. All patients received MTX at baseline and were randomized into two arms of treatment intensification if they did not reach remission after 4 months.

Set C consisted of RA-patients from the Reumazorg Zuid West Nederland (RZWN) from January 2015 till December 1st 2022 from 9 different hospitals across the south west of the Netherlands. Herein, the diagnosis of RA was defined as having an ICD-code for RA and starting with a conventional DMARD.

Across all sets, a minimum follow-up of 1-year was required to ascertain the diagnosis of RA. Prior to conducting the study, we acquired approval from the ethics committee of the LUMC. Patients and public were not involved during the development, execution, and dissemination of the study.

### Selection of phenotypic variables

To construct patient phenotypic profiles, we extracted information on serology (RF and ACPA), location of joint involvement (tender- and swollen joints (TJC and SJC)), demographics, blood profiles (hemoglobin, hematocrit, leukocyte- and thrombocytes levels) and ESR at baseline (Table S1).

Baseline was defined as the first visit to the clinic (set A&C) or the moment of inclusion in the trial (set B). Patients with missing lab or joint location variables were dropped (Fig. S2).

### Construction of the clusters

We combined the different data types from the EHR to construct a low dimensional representation of the patients (i.e. a patient embedding) with a multi-modal autoencoder (MMAE) [19]. We used PhenoGraph[26] to further organize the patients into subcommunities based on their similarities in clinical parameters. PhenoGraph was preferred over traditional methods like K-means, since it is better at handling sparse data with many variables (see the supplemental material).

### Cluster interpretation

For each cluster, we examined the characteristics and visualized the phenotype on a pictorial mannequin with an integrated heatmap to demonstrate the number of involved joints. We used a surrogate ML-technique to model the cluster assignment and subjected this model to a SHAP (SHapley Additive exPlanations) [27] analysis to retrospectively identify the most important variables per cluster. The SHAP plots show the strength and direction of impact of that variable for each patient (also those who are not assigned to that cluster).

### Cluster validation

To confirm that our identified clusters comprised a stable and relevant partitioning, we performed a number of validation checks. We examined cluster stability by measuring how often patients co-cluster across 1000 random subsets of the data and assessed possible factors influencing the partitioning. Next, we conducted a Local Inverse Simpson’s Index analysis [28] to assess whether physicians were evenly distributed across the clusters or whether there was a batch effect (see supplemental material).

To infer the clinical relevance, clinical outcomes were evaluated using a Cox regression model, including: time to MTX-failure (defined by replacement of- or adding an additional DMARD to MTX) and remission (DAS44 < 1.6) within one year. Moreover, we evaluated the replicability on an external dataset (set B & set C), where individuals were assigned to clusters in accordance with the previously learned patient embedding (see supplemental material). Finally, we checked whether the new cluster labels improved the regression model on top of the known poor prognostic factors; ACPA-positivity, RF-positivity, sex and age.

### Statistical tests

When comparing more than two groups, we used Kruskal Wallis and post-hoc Dunn’s test. For survival analysis we used the log-rank test to examine the overall trend and a univariate Cox-regression [29] to quantify the cluster differences. We inferred the DAS-remission status during the survival analysis, carrying the last observation forward if it was missing (effectively the same as time to event). The proportional hazards assumption was verified by examining the Schoenfeld residuals [30]. We examined the additive value of the clusters on top of known predictors by adjusting the Cox regression model for MTX-response on covariates. The statistical significance was inferred with ANOVA. All of our scripts are publicly available online at Github [31].

## RESULTS

### Patients

We retrieved 2,691 RA patients for training set A of whom, 1,387 were included in our study based on the availability of lab values and joint counts. For the replication, set B and set C had 364 and 1,227 RA cases, of whom 307 and 515 had complete information (Table 1, Fig. S2). Each dataset captured a typical early RA population [32, 33]. In comparison to set A, patients in the replication sets exhibited higher rates of seropositivity and had fewer tender joints. On average, set B patients were younger and set C patients had less inflammation, with a median ESR of 16. We used the phenotypic variables (see methods) of set A to construct the patient embedding.

**Table 1:**
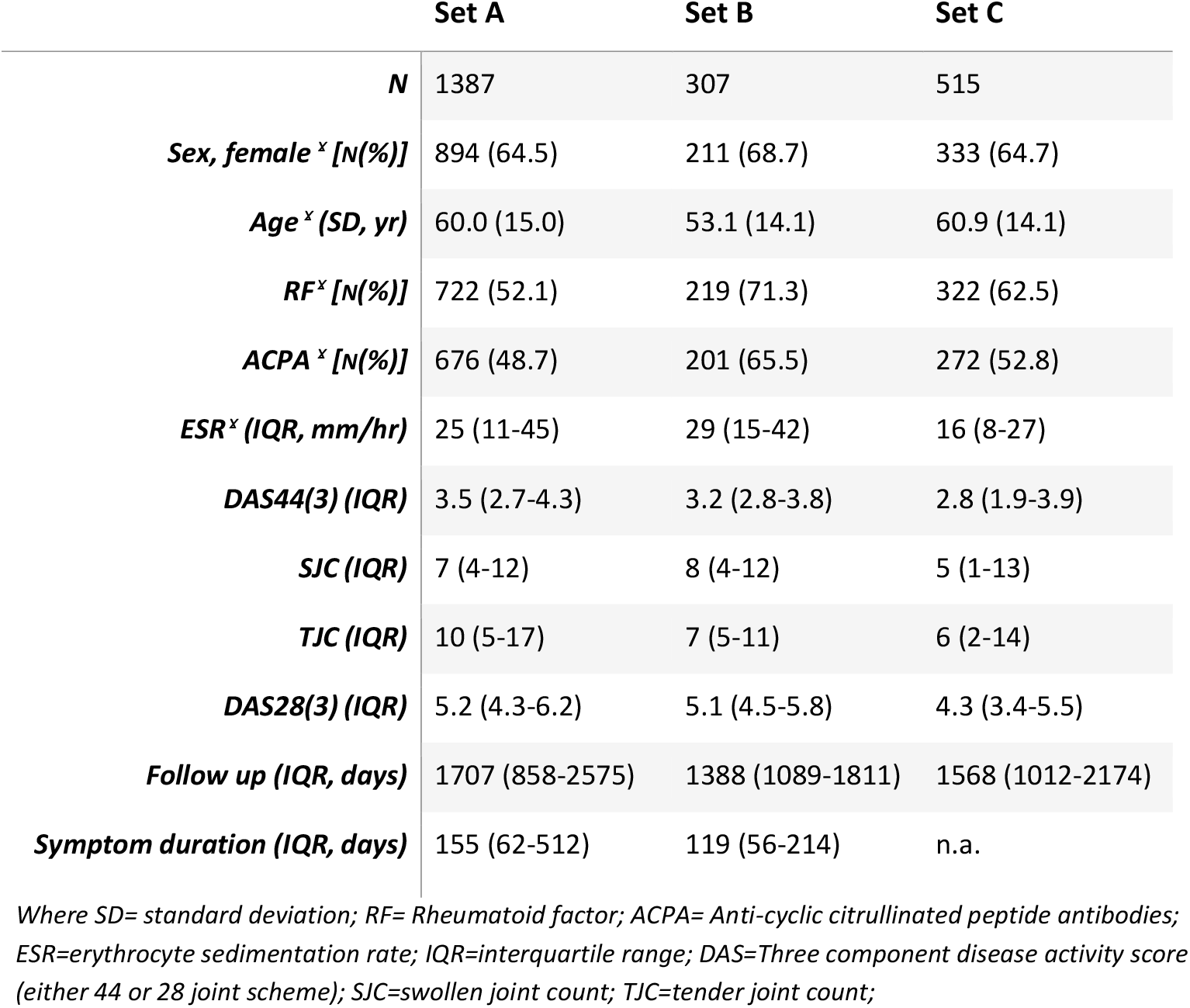
Baseline table for the development set (set A) and the replication sets (set B and C)

### Four clusters separated by joint location, serology and blood values

The patient embedding showed four different clusters (Fig. 1, S3). These clusters were not driven by any single clinical variable, as indicated by the wide dispersion of values. The clusters exhibited variations in joint location, amount of inflammation, age and differences in seropositivity (Fig. 2, Table S2):

**Figure 1:**
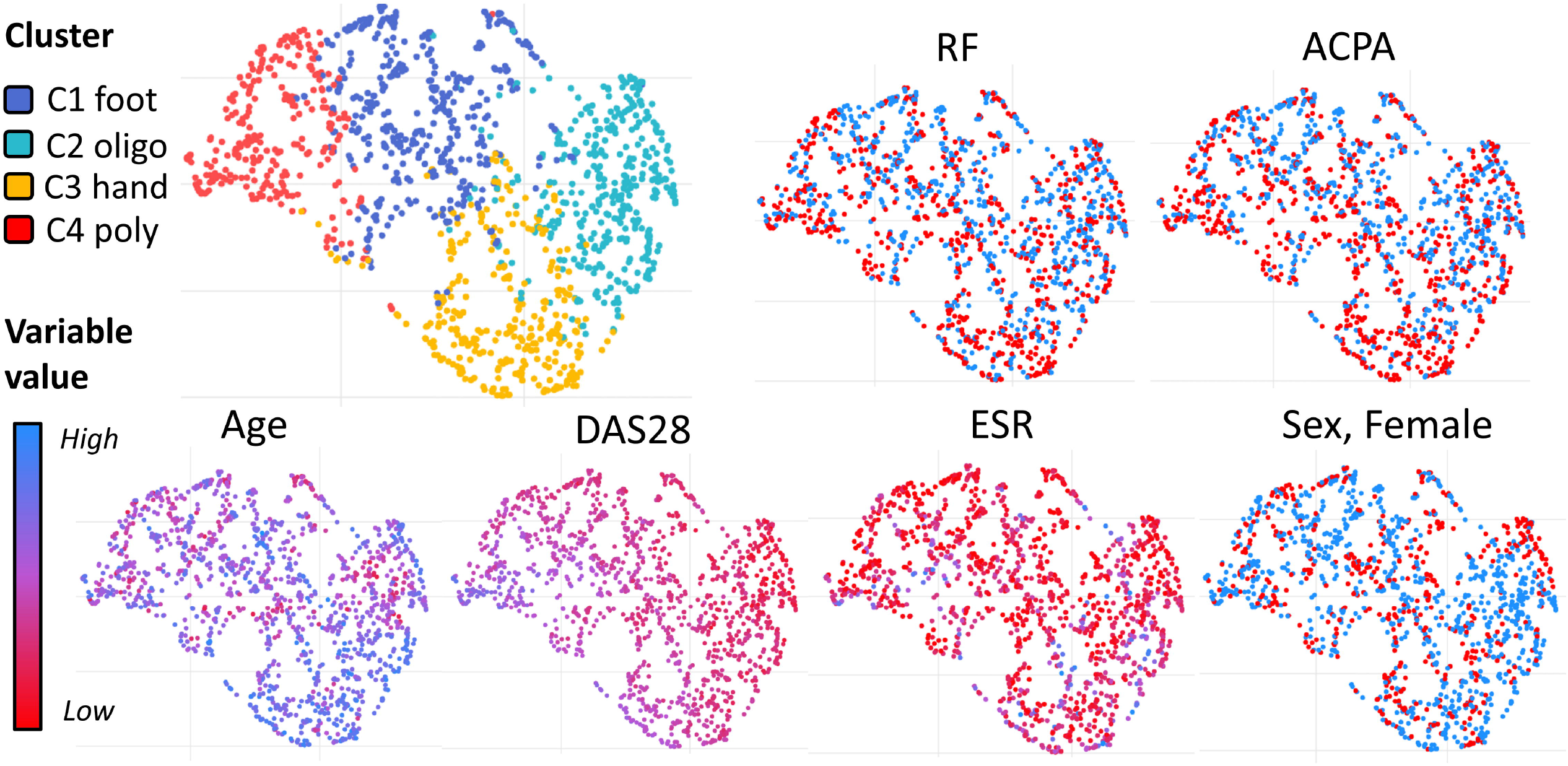
Two-dimensional Uniform Manifold Approximation and Projection (UMAP)-representation of the patient embedding. Here each patient is represented by a dot that is colored by the four clusters in the first plot and a gradient from high (blue) to low (red) in the subsequent plots. From left to right: dots are colored on corresponding cluster, Rheumatoid factor (RF) status, anti-cyclic citrullinated protein (ACPA) status, Age, disease activity score 28 (DAS28), erythrocyte sedimentation rate (ESR), Sex (1= Female, 0=Male).

**Figure 2:**
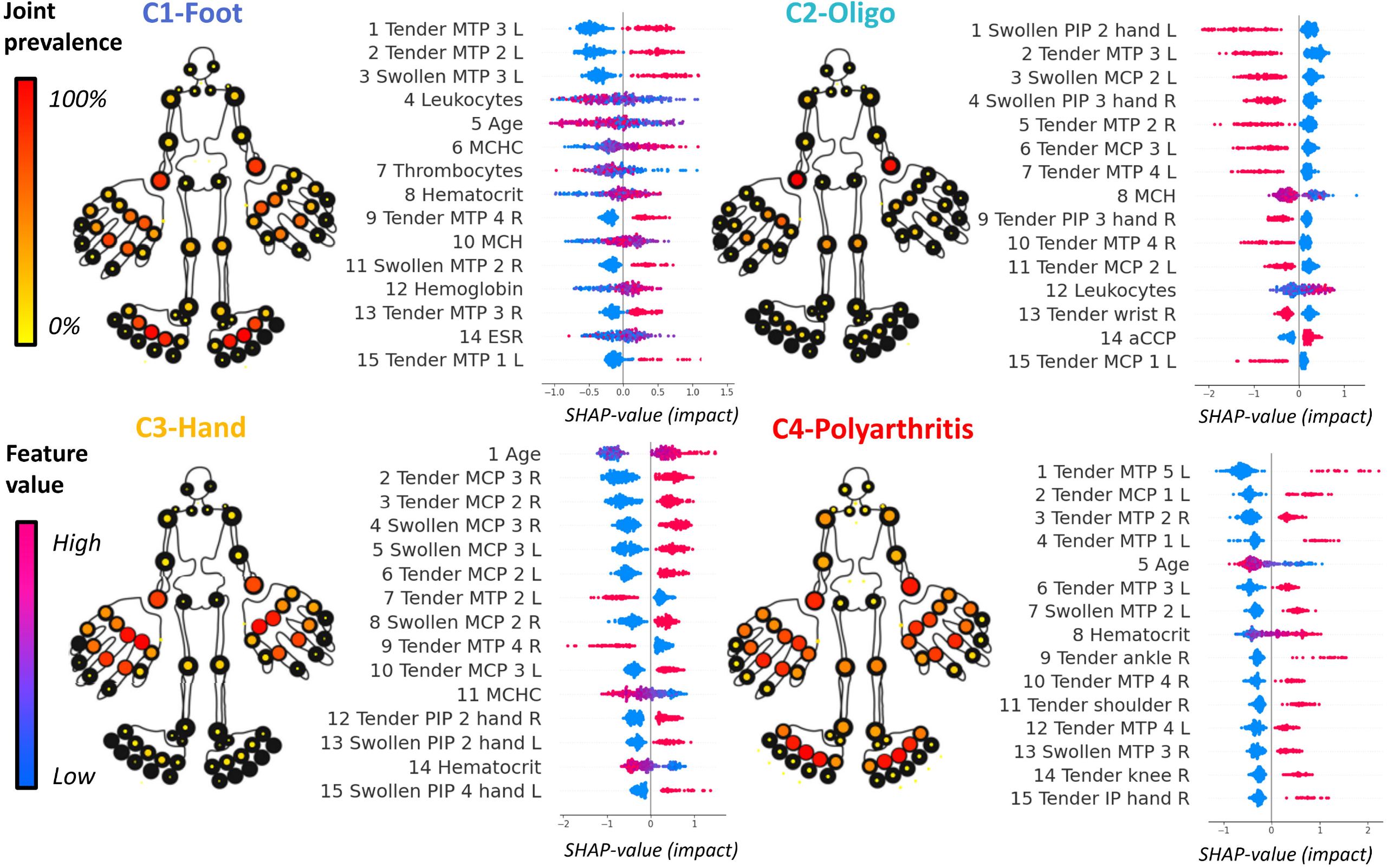
Cluster summary plots showing the average joint involvement, the top 15 driving variables. The mannequin is formatted as a heatmap, showing the prevalence (red=100%, yellow=0%) of joint involvement (tender or swollen). In the SHAP plots, the most informative features for each cluster are listed in descending order. Here the x-axis shows the strength and direction of impact of that variable for each patient (represented by a dot). The colour of the dot shows the initial value of the clinical variable (pink=high, blue=low). Where ACPA=anti-cyclic citrullinated peptide antibodies; ESR=erythrocyte sedimentation rate; IP= interphalangeal; L=left, MCH=mean corpuscular hemoglobin; MCHC=mean corpuscular hemoglobin concentration; MCP=metacarpophalangeal; MTP=metatarsophalangeal; PIP=proximal interphalangeal; R=right;

- Cluster 1 (n=415) foot

moderate number of involved joints, particularly feet joints, younger patients, low leukocyte and thrombocyte levels.

- Cluster 2 (n=380) oligo-articular

limited joint involvement and mostly seropositive patients.

- Cluster 3 (n=323) hand

elderly patients, symmetrical polyarthritis of hands, seronegative.

- Cluster 4 (n=269) polyarthritis

majority seronegative polyarthritis in hand and feet though with lower ESR.

The clusters were stable with an average >80% of patients grouping together in the same cluster over the 1000 iterations in the stability analysis (Fig. S4&S5). In fact, the stability was better in our combined multi-modal approach than if we take each data type (numeric/categorical) separately (Fig. S6). The clusters were not driven by treating physicians (Fig. S7) and were generalizable across different validation sets (Table 2, Fig. S8&S9), showing similar joint involvement patterns (Fig. 3).

**Table 2:** Baseline characteristics of the different patient clusters (set A + replication sets B & C)

|  | C1-Foot | C2-Oligo | C3-Hand | C4-Poly |
| --- | --- | --- | --- | --- |
| **N** | 596 | 761 | 450 | 402 |
| **Sex, female <sup>γ</sup> [n(%)]** | 389 (65.3) | 505 (66.4) | 272 (60.4) | 272 (67.7) |
| **Age <sup>γ</sup> (SD, yr)** | 56.6 (14.6) | 59.5 (14.6) | 66.4 (13.2) | 54.7 (14.7) |
| **RF <sup>γ</sup> [n(%)]** | 370 (62.1) | 497 (65.3) | 200 (44.4) | 196 (48.8) |
| **ACPA <sup>γ</sup> [n(%)]** | 355 (59.6) | 449 (59.0) | 162 (36.0) | 183 (45.5) |
| **ESR <sup>γ</sup> (IQR, mm/hr)** | 22 (9-36) | 24 (11-38) | 28 (13-48) | 22 (9-40) |
| **DAS44(3) (IQR)** | 3.8 (3.2-4.2) | 2.5 (2.1-3.1) | 3.6 (3.0-4.0) | 4.7 (3.7-5.4) |
| **SJC (IQR)** | 8 (5-12) | 2 (1-5) | 10 (7-15) | 15 (9-22) |
| **TJC (IQR)** | 11 (8-15) | 3 (2-5) | 11 (7-15) | 22 (15-30) |
| **DAS28(3) (IQR)** | 5.1 (4.2-5.8) | 3.9 (3.1-4.6) | 5.5 (4.8-6.2) | 6.1 (5.1-7.0) |
| **Follow up (IQR, days)** | 1307 (733-2020) | 1428 (760-2082) | 1127 (566-1875) | 1512 (1012-2246) |
| **Symptom duration \* (IQR, days)** | 143 (56-364) | 186 (70-399) | 101 (48-279) | 147 (56-357) |

Where SD, standard deviation; RF, rheumatoid factor; ACPA, anti-cyclic citrullinated peptide antibodies; ESR, erythrocyte sedimentation rate; IQR, interquartile range; DAS, three component disease activity score (either 44 or 28 joint scheme); SJC, swollen joint count; TJC, tender joint count; <sup>γ</sup>, clinical variables that were used for clustering (set A); \*, symptom duration was only calculated for patients from set A and B.

**Figure 3:**
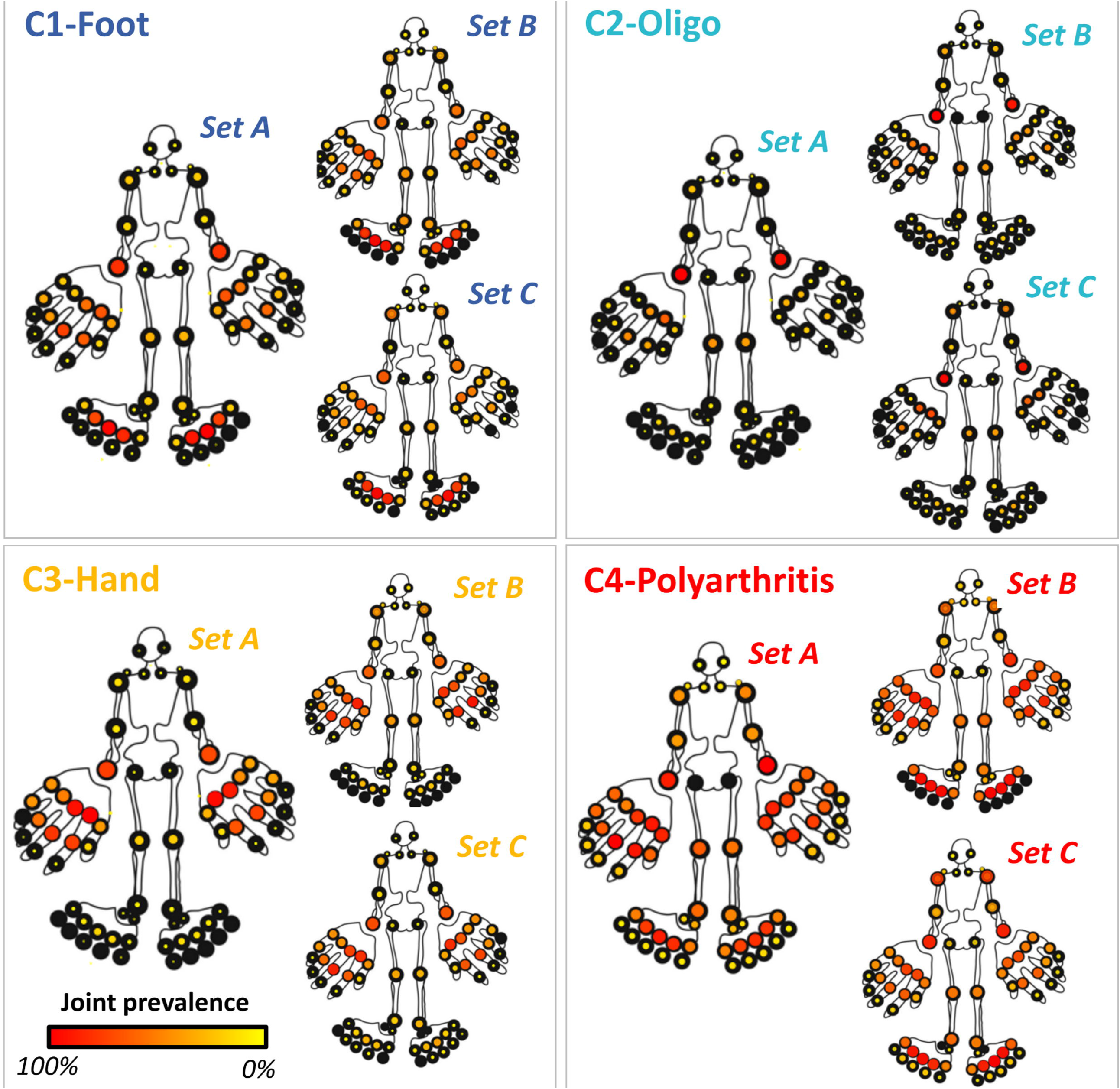
Pictorial mannequins for replication set B and C and their original counterpart (set A) to show the affected joints for each cluster with color and size to depict prevalence. Frequency is colored on a gradient from red (=100%) to yellow (=0%). When there is no colored dot, it signifies the absence of both swelling and pain at baseline for these patients.

Also, the identified patient clusters did not seem to represent different disease stages as the cluster with the longest symptom duration had the lowest joint count and vice versa. There were differences in cluster prevalence between the validation sets (Table S3&S4).

### Validation on clinical outcomes beyond baseline

In set A, 80% of patients received MTX as an initial drug across all clusters. The Kaplan Meier curves show a difference in MTX failure between the clusters: 27%, 23%, 16%, 30% (for cluster 1-4, *P*<0.001, Fig. 4a). The hand cluster (C3) had clearly the best prognosis, where patients were twice as likely to stay on MTX than the most severe disease subtype (C4) (HR 0.48 (95% CI 0.35-0.77), *P*<0.001). Additionally, the hand cluster (C3) did better than the foot cluster (C1) (HR:0.55 (0.37-0.82) *P*=0.003).

**Figure 4:**
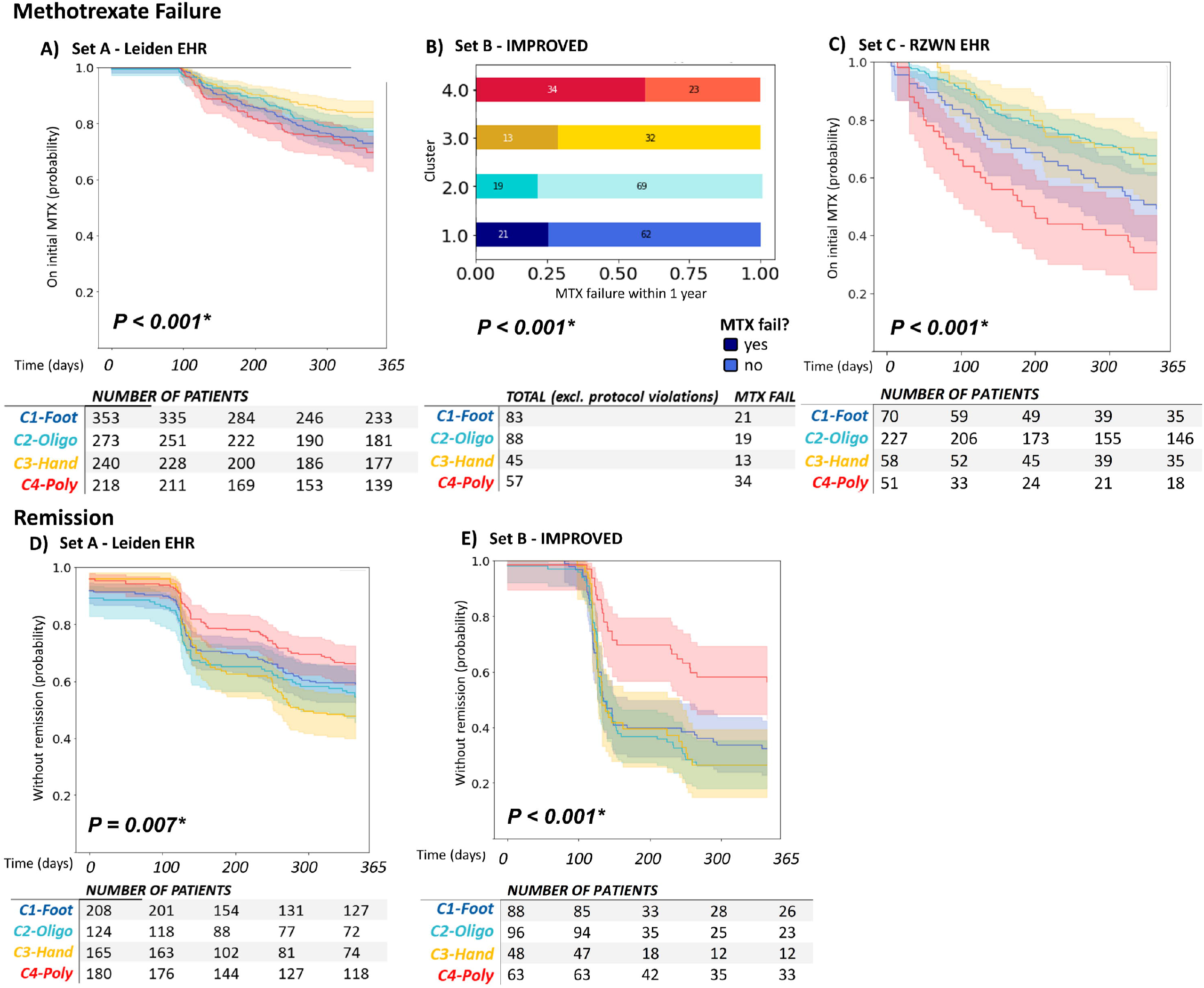
Downstream analysis depicting association of baseline clusters with methotrexate failure (A, B, C) - and remission (D, E) after 1 year. Here we generated Kaplan Meier survival curves for MTX-switch - and remission rates (DAS44<1.6) for survival time data or cross tabs for binary outcomes (i.e. set B had MTX switch integrated in protocol). Follow-up survival data is shown only for MTX-starters in set A (n=1,084; A), set B (n=273, B), set C (n=406; C) or patients with remission information in set A (n=676; D) and set B (n=295; E). Global trend was inferred with the log rank test for survival curves or chi-squared test for the cross tabs.

Consistent with MTX response, we observed differences in remission rates: 44.3%, 47.4%, 55.7%, 38.5% (for cluster 1-4, *P*=0.007, Fig. 4b) with the biggest difference between the hand C3 and polyarthritis group C4 (HR 1.65 (95% CI 1.2-2.29), *P*=0.002), also when corrected for baseline disease activity. Both the MTX and remission analyses remained significant when corrected for baseline DAS (Fig. S10).

### ACPA within the clusters

Since the literature reports that ACPA is indicative of persistent disease [34], we examined whether the ACPA status was the main factor driving the difference in MTX failure. In set A we found a higher treatment failure in ACPA positive than negative patients (27.6% versus 22.0%, Fig. 5, S11a), though it was not significant (P=0.057). Moreover, the association of ACPA with MTX-failure differed within the clusters (P<0.001, Fig. 5, S11b).

**Figure 5:**
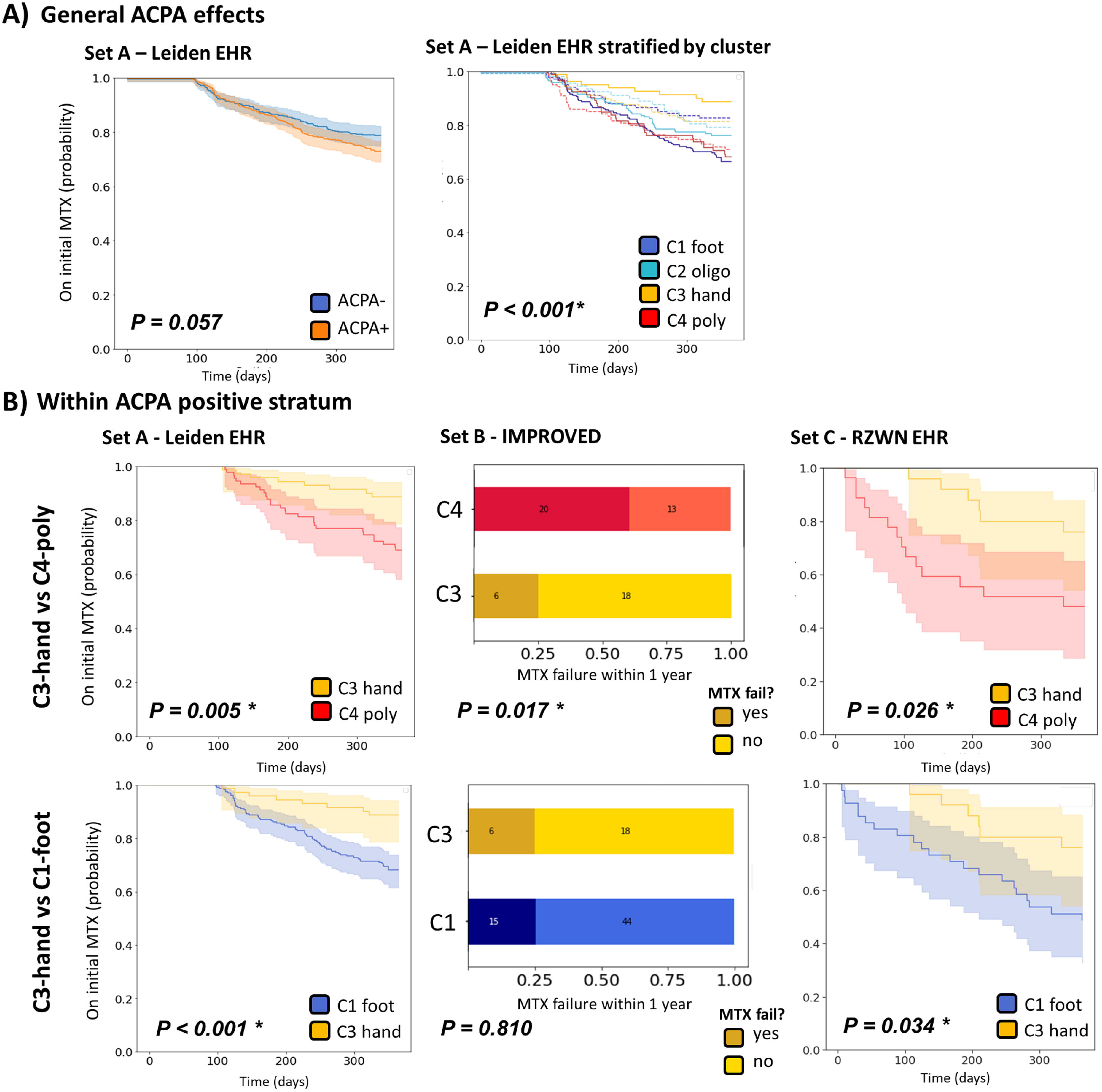
Kaplan Meier depicting the A) ACPA effect on MTX switch rates in general and stratified by cluster, B) local cluster differences between hand (C3) and foot clusters (C1 & C4) within the ACPA stratum. Here the dashed lines indicate the seropositive patients. Global trend was inferred with the log rank test for survival curves or chi-squared test for the cross tabs.

The difference between the hand-cluster C3 with the foot clusters C4 and C1 was larger within the ACPA-positive stratum (C3 vs C1 (HR:0.3 (0.15-0.60) P<0.001), C3 vs C4 HR:0.33 (0.15-0.72) P=0.005) (Fig. 5b). For remission we could not find this difference in the ACPA-positive stratum (Fig. S12a).

The good response in C3 raised the question whether this group overrepresented patients with parvovirus induced arthritis instead of RA, but none of our clusters were enriched for parvovirus positive patients (Fig. S13).

### Replication

In set C, 79% of patients received MTX as the initial drug across all clusters, whereas in set B all patients were administered MTX. In both replication sets we again observed a better outcome of C3 for MTX failure (global *P*<0.001, and *P*<0.001). Remission could only be tested in replication set B where it confirmed our previous finding (*P*<0.001). Consistent with the original finding, the difference between C3 and C4 was particularly strong in the ACPA positive stratum in both replication set B (OR: 0.22 (0.12-0.63) *P*=0.017) and set C (HR:0.38 (0.22-0.68) *P*<0.001). Within the ACPA positive stratum we also found the significant difference between C3 and C1 back in replication set C (HR:0.37 (0.15-0.93), *P*=0.034), though not in replication set B (OR: 1.03 (0.34-3.05) *P*=0.801). All the analyses remained significant when corrected for baseline DAS (Fig. S14b-c).

### Informative value of clusters beyond known risk factors for MTX failure

To ascertain the additive value of our clusters, we added the cluster information to a Cox regression model of baseline variables known to associate with MTX failure (Fig. S15). Here, the addition of the clusters as a covariate significantly improved the fit of the model (*P*=0.013). The inclusion of other well-known contributing factors like differences in disease or symptom duration, delay in treatment or the number of affected joints (Table S5) did not diminish the additive value of clustering.

When we encoded the joint location into two binary variables for a) foot involvement and b) hand involvement and added this to the initial model, the clusters no longer improved the model on MTX-failure. Thus, the hand and feet count provided a proxy for cluster membership in this context (Fig. S16).

## DISCUSSION

Through the application of deep learning and clustering, we identified four phenotypically distinct subtypes of RA at baseline visit. These were characterized by arthritis in feet (C1), seropositive oligoarticular disease (C2), seronegative arthritis in hands (C3) and polyarthritis (C4). We obtained these results using unsupervised pattern recognition on real world clinical data. Compared to the traditional, hypothesis based analysis, this approach has the potential to identify novel non-linear clinical signatures that tend to be overlooked. On the other side there is a risk of finding meaningless structure that is driven by noise or bias in the data. Therefore, we rigorously tested the credibility of our findings: demonstrating that our clusters are stable and not influenced by physicians or disease stage. Next, we validated the relevance of our clusters with one-year clinical outcomes and replicated both the clusters and clinical outcomes in two separate datasets.

We noticed a clear difference in treatment success between the hand and foot clusters. Both foot clusters (C1,C4) showed higher MTX failure and less remission rates compared to the hand cluster (C3). Intriguingly, this difference in outcome was not explained by differences in number of involved joints nor symptom duration at baseline or initiation of treatment. The poor prognosis for inflamed feet was already suggested in a previous cross-sectional study [35]. In this study, researchers noted a predominance of feet/ankle-involvement for patients that remained having active disease under treatment. However, this study did not analyze these differences in an untreated baseline population. To our knowledge we are the first to show that feet involvement at disease presentation is associated with less treatment success and that this is independent of disease activity or symptom duration. The risk of treatment failure in those with foot involvement was similarly bad as the presence of ACPA-positivity and is a risk factor independent of ACPA.

This observation is relevant as foot complaints are quite common [35], yet many disease activity scores exclude the joints in the lower extremities. The problem of this was previously noted by patients and rheumatologists who called for methods that consider the lower extremities [36]. Our study further supports this notion.

Interestingly, within the ACPA-positive stratum the difference in MTX success between the hand and foot clusters was strongest. Notably, in all replication sets we found a significant difference between the hand-cluster C3 versus the poly-cluster C4, and in 2 out of 3 datasets we also found an increase compared to the foot cluster of C1. Suggesting that arthritis in the feet has a worse response to MTX than arthritis in the hands. For remission we did not observe a stronger effect in the ACPA positive stratum for remission rates. This could be a result of the success of target therapies that suggest intensifying treatment rapidly, particularly for ACPA-positive patients.

There are several possible explanations for the good outcome of patients in the hand cluster (C3). As shown, it is not explained by symptom duration or disease activity at baseline nor the lower prevalence of ACPA in that cluster. The short symptom duration combined with a high number of inflamed joints and low prevalence of seropositivity gives the impression of a self-limiting reactive arthritis[37,38]. We checked for parvo-virus positivity and did not find any difference in parvovirus positivity between the clusters. Still, C3 may depict a form of naturally more self-remitting disease and further research into this may provide novel treatment insights. An alternative option is that current drugs are selected on being successful for the C3 like patients, as drug trials frequently used outcome measures that ignore the lower extremities. Unfortunately, our study lacks power to test any treatment association beyond methotrexate. Clearly, further studies are needed to test these hypotheses.

As a positive control, we looked for age related subsets. Consistent with previous literature, our clusters captured the elderly onset RA (EORA) cluster (C3) as previously described: casting higher inflammation (ESR and SJC), lower prevalence of women and lower prevalence of ACPA and RF compared to the total group of RA patients.[9, 10] Furthermore, our study uncovers more granular subtypes than just the distinction between EORA and young onset RA (YORA). There are younger people with a clinical pattern similar to the EORA in C3 and older people that cluster with younger seropositive patients.

In contrast with the general assumption that ACPA divides RA into two sub entities, we did not find a clear ACPA dichotomy. Notably, previous studies that examine the disease at baseline do not find clear differences based on ACPA status either, as this was only observed in established disease[34,39]. We found that the prevalence of ACPA was lowest in the more RA typical clusters with polyarthritis (C3 and C4) and highest in the cluster with the lowest joint involvement (C2). In set B and set C we observed a similar pattern of ACPA prevalence between the clusters as in set A though less prominent.

The higher prevalence of ACPA in the oligo-articular subset could be a result of the RA classification criteria allowing an RA diagnosis when people have polyarthritis or are ACPA positive.[40] We aimed to reduce this effect by using the RA diagnosis over the course of a year, which overcomes most of the initial clinical misclassification. Also, for set A and C, we did not use the classification criteria, but the diagnosis of the physician instead.[24] Encouragingly, the findings were consistent between the three datasets. Our finding of C1 with high ACPA-positivity prevalence as well as a high number of inflamed joints also underlines that the clusters were not (completely) driven by criteria induced misclassification.

Notably, this higher ACPA-positivity in the foot cluster-1 confirms the recent finding of the Leiden clinically suspect arthralgia cohort of an increased prevalence of feet in the ACPA positive RA population compared to the ACPA negative patients [41].

There are some limitations to our study that merit closer inspection. First, we had to define MTX success based on adding or switching to a different drug. Probably some of the switches were due to side effects unrelated to efficacy. We do not expect this has led to false-positive results, as side effects are more likely to have been present in the elderly cluster (C3) where we observed the opposite, namely less treatment switches. Secondly, the therapeutic approach likely differs from center to center, which may affect the outcome. For example, in the outpatient clinic of Leiden, rheumatologists appear to be less likely to switch at an early stage, whereas we found that in RZWN many switches already occurred in the first few months. These differences did not lead to a difference in the association of clusters with treatment success.

Our study does not test the stability of the clusters over time. Previous literature describes the recurring presence of inflammation in identical joints over the course of the disease [42]. When considered alongside the observation that our subsets are partitioned based on joint involvement patterns, it suggests our clusters will be stable over time. This is also supported by the previously mentioned study that showed cross-sectionally that foot inflammation was associated with a decreased change of remission[35].

Important to underline is that our identified clusters are not set in stone. Though we observed a high robustness of our clusters, patients laid on a gradient (Fig. 2) and did not segregate in clearly separable modules. The cluster structure that we identified could also be summarized into more or fewer clusters and the clusters might become clearer when more layers of information are added. Such types of information could be genetics, gene expression patterns and molecular profiles from blood and synovial biopsies.[14, 43].

Within these limitations, we demonstrated the power of unsupervised and data-driven tools to unlock hidden structures in the data. The integration of different EHR-modalities resulted in more stable clusters than clustering on categorical or numerical variables alone. In particular, the inclusion of joint involvement patterns appears to be a major axis of variation.

In conclusion, by clustering RA patients on their first presenting symptoms, we uncovered four baseline phenotypes characterized by hand and feet involvement that are associated with one-year clinical outcomes. Our data-driven approach offers a more granular picture of RA in the clinic than the dichotomous division by age or ACPA alone. The cluster differences in outcome and clinical presentation may be indicative of a distinct etiology. Inevitably, this necessitates future research to affirm a potential biological link.

## Data Availability

We have made our scripts available in a public repository at: https://github.com/levrex/EHR-Clustering-RA. Study data is available upon reasonable request.

## FOOTNOTES

### Contributors

RK developed the study design together with EB, TM and MP. TM ran the cluster analysis, while SB ran the permutation analysis. BBdK repeated the survival analysis in the replication set. SAB and CA provided the replication data of the IMPROVED trial, while KG and JVvD provided the data from Reumazorg Zuid West Nederland. All authors contributed to the interpretation of the results, and provided ideas for further downstream analysis. AHM annotated the classification criteria. RK and TM drafted the first version of the manuscript. All authors reviewed and approved the final draft of the manuscript to be submitted.

## Patient and public involvement

Patients and/or the public were not involved in the design, or conduct, or reporting, or dissemination plans of this research.

## Ethics approval

We received ethical approval from the Medical Ethics Committee (METC) at Leiden University Medical Center according to study protocol B18.057.

## Acknowledgements

We want to thank Samantha Jurado-Zapata and David Steeman for their help with respect to the extraction and processing of the Electronic Health Record data from the Leiden University Medical Center. In addition, we want to extend our thanks to Bas van der Wal for preparing the Reumazorg Zuidwest Nederland data and Joy van der Pol for preparing the IMPROVED trial data.

