## supplemental material for "Location of joint involvement differentiates rheumatoid arthritis into different clinical subsets"

**METHODS**

In this document we will provide further detail on the approach that was taken for this data-driven analysis. For our study we used hospital data from Leiden (set A), the historic IMPROVED trial (set B) and hospital data from Reumazorg Zuid West Nederland (RZWN, set C). We have created a technical workflow to outline the various steps employed in this study (Fig. S17).

*Patients*

We required at least 1 year of follow-up to ensure certainty about the RA diagnosis. In set A, RA cases were retrieved using our previously developed algorithm that identifies the RA diagnosis from the format-free conclusion section of the physician notes written by the treating rheumatologist. Hereby, we only considered patients after the initiation of the digital EHR-system on August 29th, 2011, till the most recent data freeze on December 1st 2022. We used a balanced cut-off of 0.83 which corresponded to a sensitivity and positive predictive value of 0.85 [1, 2].

For set B, we used the IMPROVED trial as a historic validation set as it ran from March 2007 till 2010. The IMPROVED trial included DMARD naive RA and undifferentiated arthritis (UA) patients. In this trial patients were by default subjected to MTX treatment with tapered prednisone and randomised to different methotrexate treatment arms after 4 months if they had not reached remission by then. Here, RA cases were identified by meeting the 2010 RA criteria[3].

Set C comprised patients treated at Reumazorg Zuid West Nederland (RZWN) from January 2015 until December 1st, 2022, spanning nine hospitals across the southwest region of the Netherlands. RA patients were selected if they featured a specific ICD code ('M06.99', 'M06.09', 'M05.99', 'M06.90', 'M05.29', 'M05.19+') and if they started with csDMARD therapy.

*Data preprocessing*

In order to optimally integrate the different data modalities of our EHR data, we split the data into two layers: 1) numerical (hematology workup, demographics) and 2) categorical variables (binary serological markers (ACPA/RF) and location of joint inflammation and tenderness). The numerical modality underwent a Yeo-Johnson transformation to cast each variable to a normal distribution, with the exception of the log-normally distributed serum ESR levels, which were log-transformed. The categorical variables were subjected to one-hot-encoding, resulting in a binary variable (present/absent) per category of each variable.

*Construction of patient embedding*

After preprocessing, we used a variational autoencoder technique called Multi Modal Auto Encoder (MMAE) [4] to construct a patient embedding. A patient embedding is a condensed representation of patient-related data. This representation is a combination of latent factors that capture most variation in the data. These embeddings can be used to cluster patients, identify patterns, build predictive models or assist in making disease classifications.

MMAE has the added benefit that it can work with different modalities. Autoencoders are typically used to reduce the dimensionality of the data by training a network of non-linear relationships between variables. In effect, this not only removes the redundant information between variables, but also has a denoising effect. To exclude the possibility that our model was overfitting we checked for a loss in quality between the patient embedding of the training and validation set and examined the contribution of each EHR-layer to the eventual integrated space with a flameplot.[5] Next, the patient embedding was truncated into a two-dimensional space with UMAP.[6] Similar to MMAE, UMAP is recognized for its non-linear capabilities.

*Defining the optimal value of k*

For clustering we use Phenograph, a graph based clustering algorithm, that groups patients together based on their clinical commonalities. The distance between any two patients is expressed with the Minkowski metric.

PhenoGraph utilizes k nearest neighbours to construct the graph and the Louvain algorithm [7] for community detection. To define k, we tested different configurations for our clustering: by comparing the similarity across the different clustering configurations. In the heatmap we notice that there are multiple stable 'chunks' in which we achieve relatively robust clusters. Looking at the parametric space, we ultimately chose a k of 270, as it captured most variation while still being stable (Fig. S3). The k of 270 corresponded to four clusters.

*Cluster validation*

To ensure that our identified phenotypic clusters did not capture random irrelevant patterns but stable and meaningful patterns instead, we performed several validation checks. We evaluated the stability of clusters across different parametric configurations of the algorithm and across the subsample space by conducting a (1000 times) bootstrapping analysis. This procedure consists of taking multiple random subsets of the data to analyze the stability to evaluate the likelihood of patients ending up in the same cluster. We compared the cluster stability between our combined modality approach to an approach using only a single modality.

Furthermore, we investigated the possibility that our clusters were biased by the treating physician. We used the Local Inverse Simpson's index (LISI) [8] to quantify the local diversity of physicians across the clusters. The LISI effectively calculates the sum of unique physicians in the local neighbourhood of a patient (which is 15 neighbors by default) within a cluster. We rendered a density plot to depict the distribution per cluster. This figure was complemented by a stacked bar chart to visualize the occurrence of physicians.

*SHAP analysis: Identifying the driving features per cluster*
To identify the driving features for each cluster we performed a SHAP analysis.[9] This is a technique borrowed from game theory, where it is used to estimate the contribution
of each participant to a victorious outcome. In other words, SHAP is used to calculate their contribution to the total gain (surplus). SHAP is considered an agnostic method, since the impact of each feature (i.e. the SHAP value) has to be estimated in a reverse-engineering manner, as it cannot be retrieved from the intrinsic structure. Nowadays, SHAP is commonly used to render increased insight into deep learning techniques.

In the present study, we use SHAP to estimate the relative impact of a variable to the cluster assignment per patient. We trained a surrogate model (XGBoost classifier) to predict cluster membership to identify the most important variables for each cluster. Next, we evaluated the quality of this model in an independent test set via a confusion matrix.

*Survival analysis*
We checked whether the clusters corresponded to clinical outcomes after one year, namely: MTX-failure and remission. Starting with MTX, we defined treatment failure as a treatment switch: either changing or complementing MTX with a different drug. We took the date of the first treatment prescription as a baseline (t=0), to guarantee that our results weren't biased by the fact that certain patients start treatment way earlier than other patients

Schoenfeld residual plots were produced to test the proportional hazards assumption [10]. We fit a LOWESS (Locally Weighted Scatterplot Smoothing) line on the Schoenfeld residuals to check whether the proportional hazards assumption of the Cox-regression is met.

*Validate on clinical outcomes beyond baseline*
Next, we checked if our subclusters associate with clinical outcomes within one year, namely: time to MTX-failure (defined by replacement of- or adding a DMARD to MTX) and remission (DAS44 < 1.6). In addition, we checked if the MTX-association was a result of chance by testing the MTX response association for randomly assigned clusters for 1000x iterations.

To ensure our cluster difference in MTX-outcome was not merely driven by already known clinical markers associated with response (RF, ACPA, Age, sex, joint counts [11-13]), we trained a Cox-regression model with- and without clusters. The additive value of our clusters was established by comparing the two Cox models with an ANOVA based on Chi-squared. Moreover, we fitted another Cox-regression model to make sure that the MTX-association was not mediated by differences in symptom duration, delay in treatment and number of affected joints.

*Independent validation in replication set*
Finally, we performed an external validation in historic trial data (set B) and data from another center (set C) to test if our clusters were recurring across the different datasets. Clustering techniques generally require recomputing the clusters altogether when novel patients are added. Since we wanted to map novel instances to our discovered clusters we build POODLE (**P**rojecting **O**bservations **O**n a **D**eep **L**earned **E**mbedding) [14]. We trained and tested POODLE on separate subsets of the original development data (Set A).

Next, we projected the novel patients and examined their distribution in the new space. The clustered replication patients were compared to the original clusters by examining their characteristics, joint inflammation patterns and SHAP-profiles. To establish the generalizability of our clusters, we repeated the survival analysis for MTX-failure and remission in the external data.

Due to the trial design of set B, patients switch automatically from the initial therapy if they don't reach remission by 4 months. Therefore, we treated MTX-failure in set B as a binary outcome rather than time-to-event. Given the observation that ACPA-positivity is associated with a more severe prognosis, we also checked if the cluster differences increased within the ACPA-positive stratum.

To ensure that the cluster difference in outcome was not solely attributed by baseline DAS44 alone, we fitted two Cox-regression models for time till MTX switch within one year: a reduced model with baseline DAS44 and a full model comprising both DAS44 and the clusters. The additive value of the clusters was inferred by comparing the two models with an ANOVA based on Chi-squared. For set B we used a Logit regression model instead of a Cox-regression since we had a fixed time till MTX-switch.

*Power analysis*
Before conducting the survival analysis, we tested the minimal required samples under Cox Proportional Hazard [15]. The postulated hazard ratio & remission rates were based on the means of the biggest cluster difference in survival curves after 1 year in the Leiden data (set A). We rendered a power curve to show the correspondence between the number of patients and corresponding statistical power, requiring a minimum of 0.8 to proceed with the replication.

### RESULTS

*Patient*
In set A and set C we dropped patients with missing EHR-information. To ensure that our learned patient embedding was not biased towards a specific subpopulation of RA, we checked for any differences between the patients with complete and incomplete electronic patient dossiers. Herein, we did not find any differences for set A or set C (Table S6, S7). We noticed that the ESR, Thrombocytes, Leukocytes and the physical examination (mannequin) values were most often missing (Fig. S18).

*Construction of patient embedding*
We used MMAE to integrate different EHR-layers into a patient embedding. Since autoencoders are known to overfit when dealing with limited samples, we checked the reconstruction loss for both training and validation set. Reassuringly, the reconstruction loss for the training- and validation set was similar (Fig. S19).

Next, we applied clustering on the patient embedding and identified 4 different clusters (Fig. 1A). The categorical variables informed the clusters to a larger extent (40%) than the numerical variables (32%) (Fig. S20).

*Cluster validation*
The clusters were highly stable as >80% of patients co-clustered (Fig. S4-S6) together across 1000 bootstrapped iterations on random subsets of the data. Moreover, the clusters were not biased by physicians as indicated by LISI plot (Fig. S7), which shows that they generally had the same distribution and were surrounded by 9 neighboring physicians on average. Furthermore the stacked bar chart demonstrated that all of the clusters had a similar palette of physicians.

Cluster 2 had a longer symptom duration (Fig. S21) compared to the other clusters, which is likely due to the fact that these patients can afford such a long duration due to relatively low joint involvement. Surprisingly, the polyarthritis group (C4) had the longest delay in time till treatment (Fig. S22), despite being the group with the most joint involvement.

*SHAP analysis*
To identify the most important variables, we performed a SHAP analysis based on a surrogate XGB model. The post-hoc XGB model assigned patients to the correct cluster in 78% of the cases in a hold-out of set A (Fig. S23).

*Survival analysis*
Based on the Cox-regression we find a significant difference between MTX-failure and the clusters, that could not be explained by other clinical variables (Fig. S16). Patients from C3-hand were more likely to stay on MTX, as compared to patients from C4-polyarthritis and C1-foot (Fig. 5). Within the ACPA-stratum, the effect size became even larger (Fig. S14). Notably, the association between MTX-response and our clusters was not just a random fluke since the p-value with our clusters was lower compared to random cluster assignment in 99.9% of the instances (Fig. S24).

Moreover, we found that proportional hazards assumption was not violated since the fitted LOWESS line was stable for most of the time (Fig. S25; rank-transformed $p=0.4204$; km-transformed $P=0.3900$).

*Independent validation in replication set*
To detect our clusters in novel data of set B and C, we built POODLE. POODLE was trained and evaluated on representative subsets of the original development set (set A), where it achieved a high accuracy of 99% for assigning patients to the right cluster (Fig. S26).

Projecting set B and set C onto the embedding obtained with set A, showed that the patients were distributed across all the clusters (Fig. S27, S28 and Table S3, S4). This demonstrates that our clusters are not dataset dependent and can be found back in historic and external data. Both the Mannequin gestalt and SHAP figures (Fig. 3, S8, S9) indicate that the palette of each cluster is well preserved (a.k.a. it constitutes the same subsets of patients): feet, few joints, hand and severe polyarthritis.

Similar to set A, we find a significant global trend between clusters and clinical outcomes after 1 year (Fig 4). Moreover, the cluster difference in MTX and remission between C3 and C4 is consistently present across the different replication sets.

*Power analysis*
Prior to replicating the survival analysis in the external dataset from RZWN we conducted a power analysis (Fig. S29). Here, the postulated hazard ratio was defined according to the biggest cluster difference (C3 and C4) for remission and MTX failure (HR 1.65 (95% CI 1.2-2.29). Based on the power curves we concluded that we had sufficient data to detect the MTX difference in RZWN (power=0.91), but not enough to detect the difference in remission rate (power=0.36).
