## supplemental tables for "Location of joint involvement differentiates rheumatoid arthritis into different clinical subsets"

***Table S1. All variables per layer in the Electronic Health Records used for clustering set A and the replication sets B and C (*= missing for both B&C, **= only missing for set C)***

| LAYER | COLUMNS | |
|---|---|---|
| MANNEQUIN_OHE | Swollen_elbow_L_neg | Swollen_elbow_L_pos |
| | Swollen_IP_hand_L_neg | Swollen_IP_hand_L_pos |
| | Swollen_IP_hand_R_neg | Swollen_IP_hand_R_pos |
| | Swollen_IP_foot_L_neg | Swollen_IP_foot_L_pos |
| | Swollen_IP_foot_R_neg | Swollen_IP_foot_R_pos |
| | Swollen_acromioclaviculair_L_neg | Swollen_acromioclaviculair_L_pos |
| | Swollen_acromioclaviculair_R_neg | Swollen_acromioclaviculair_R_pos |
| | Swollen_ankle_L_neg | Swollen_ankle_L_pos |
| | Swollen_ankle_R_neg | Swollen_ankle_R_pos |
| | Swollen_cervical spine_neg | Swollen_cervical spine_pos |
| | Swollen_CMC_L_neg | Swollen_CMC_L_pos |
| | Swollen_CMC_R_neg | Swollen_CMC_R_pos |
| | Swollen_DIP_2_hand_L_neg | Swollen_DIP_2_hand_L_pos |
| | Swollen_DIP_2_foot_L_neg | Swollen_DIP_2_foot_L_pos |
| | Swollen_DIP_2_hand_R_neg | Swollen_DIP_2_hand_R_pos |
| | Swollen_DIP_2_foot_R_neg | Swollen_DIP_2_foot_R_pos |
| | Swollen_DIP_3_hand_L_neg | Swollen_DIP_3_hand_L_pos |
| | Swollen_DIP_3_foot_L_neg | Swollen_DIP_3_foot_L_pos |
| | Swollen_DIP_3_hand_R_neg | Swollen_DIP_3_hand_R_pos |
| | Swollen_DIP_3_foot_R_neg | Swollen_DIP_3_foot_R_pos |
| | Swollen_DIP_4_hand_L_neg | Swollen_DIP_4_hand_L_pos |
| | Swollen_DIP_4_foot_L_neg | Swollen_DIP_4_foot_L_pos |
| | Swollen_DIP_4_hand_R_neg | Swollen_DIP_4_hand_R_pos |
| | Swollen_DIP_4_foot_R_neg | Swollen_DIP_4_foot_R_pos |
| | Swollen_DIP_5_hand_L_neg | Swollen_DIP_5_hand_L_pos |
| | Swollen_DIP_5_foot_L_neg | Swollen_DIP_5_foot_L_pos |
| | Swollen_DIP_5_hand_R_neg | Swollen_DIP_5_hand_R_pos |
| | Swollen_DIP_5_foot_R_neg | Swollen_DIP_5_foot_R_pos |
| | Swollen_elbow_R_neg | Swollen_elbow_R_pos |
| | Swollen_hip_L_neg | Swollen_hip_L_pos |
| | Swollen_hip_R_neg | Swollen_hip_R_pos |
| | Swollen_knee_L_neg | Swollen_knee_L_pos |
| | Swollen_knee_R_neg | Swollen_knee_R_pos |
| | Swollen_MCP_1_L_neg | Swollen_MCP_1_L_pos |

| | | |
|---|---|---|
| | Swollen_MCP_1_R_neg | Swollen_MCP_1_R_pos |
| | Swollen_MCP_2_L_neg | Swollen_MCP_2_L_pos |
| | Swollen_MCP_2_R_neg | Swollen_MCP_2_R_pos |
| | Swollen_MCP_3_L_neg | Swollen_MCP_3_L_pos |
| | Swollen_MCP_3_R_neg | Swollen_MCP_3_R_pos |
| | Swollen_MCP_4_L_neg | Swollen_MCP_4_L_pos |
| | Swollen_MCP_4_R_neg | Swollen_MCP_4_R_pos |
| | Swollen_MCP_5_L_neg | Swollen_MCP_5_L_pos |
| | Swollen_MCP_5_R_neg | Swollen_MCP_5_R_pos |
| | Swollen_MTP_1_L_neg | Swollen_MTP_1_L_pos |
| | Swollen_MTP_1_R_neg | Swollen_MTP_1_R_pos |
| | Swollen_MTP_2_L_neg | Swollen_MTP_2_L_pos |
| | Swollen_MTP_2_R_neg | Swollen_MTP_2_R_pos |
| | Swollen_MTP_3_L_neg | Swollen_MTP_3_L_pos |
| | Swollen_MTP_3_R_neg | Swollen_MTP_3_R_pos |
| | Swollen_MTP_4_L_neg | Swollen_MTP_4_L_pos |
| | Swollen_MTP_4_R_neg | Swollen_MTP_4_R_pos |
| | Swollen_MTP_5_L_neg | Swollen_MTP_5_L_pos |
| | Swollen_MTP_5_R_neg | Swollen_MTP_5_R_pos |
| | Swollen_talo-calcaneo-navicularis_L_neg | Swollen_talo-calcaneo-navicularis_L_pos |
| | Swollen_talo-calcaneo-navicularis_R_neg | Swollen_talo-calcaneo-navicularis_R_pos |
| | Swollen_PIP_2_hand_L_neg | Swollen_PIP_2_hand_L_pos |
| | Swollen_PIP_2_foot_L_neg | Swollen_PIP_2_foot_L_pos |
| | Swollen_PIP_2_hand_R_neg | Swollen_PIP_2_hand_R_pos |
| | Swollen_PIP_2_foot_R_neg | Swollen_PIP_2_foot_R_pos |
| | Swollen_PIP_3_hand_L_neg | Swollen_PIP_3_hand_L_pos |
| | Swollen_PIP_3_foot_L_neg | Swollen_PIP_3_foot_L_pos |
| | Swollen_PIP_3_hand_R_neg | Swollen_PIP_3_hand_R_pos |
| | Swollen_PIP_3_foot_R_neg | Swollen_PIP_3_foot_R_pos |
| | Swollen_PIP_4_hand_L_neg | Swollen_PIP_4_hand_L_pos |
| | Swollen_PIP_4_foot_L_neg | Swollen_PIP_4_foot_L_pos |
| | Swollen_PIP_4_hand_R_neg | Swollen_PIP_4_hand_R_pos |
| | Swollen_PIP_4_foot_R_neg | Swollen_PIP_4_foot_R_pos |
| | Swollen_PIP_5_hand_L_neg | Swollen_PIP_5_hand_L_pos |
| | Swollen_PIP_5_foot_L_neg | Swollen_PIP_5_foot_L_pos |
| | Swollen_PIP_5_hand_R_neg | Swollen_PIP_5_hand_R_pos |
| | Swollen_PIP_5_foot_R_neg | Swollen_PIP_5_foot_R_pos |
| | Swollen_wrist_L_neg | Swollen_wrist_L_pos |
| | Swollen_wrist_R_neg | Swollen_wrist_R_pos |
| | Swollen_shoulder_L_neg | Swollen_shoulder_L_pos |
| | Swollen_shoulder_R_neg | Swollen_shoulder_R_pos |
| | Swollen_sternoclavicular_L_neg | Swollen_sternoclavicular_L_pos |
| | Swollen_sternoclavicular_R_neg | Swollen_sternoclavicular_R_pos |
| | Swollen_tarsometatarsal_L_neg | Swollen_tarsometatarsal_L_pos |
| | Swollen_tarsometatarsal_R_neg | Swollen_tarsometatarsal_R_pos |
| | Swollen_temporomandibular_L_neg | Swollen_temporomandibular_L_pos |

| | | |
|---|---|---|
| | Swollen_temporomandibular_R_neg | Swollen_temporomandibular_R_pos |
| | Tender_elbow_L_neg | Tender_elbow_L_pos |
| | Tender_IP_hand_L_neg | Tender_IP_hand_L_pos |
| | Tender_IP_hand_R_neg | Tender_IP_hand_R_pos |
| | Tender_IP_foot_L_neg | Tender_IP_foot_L_pos |
| | Tender_IP_foot_R_neg | Tender_IP_foot_R_pos |
| | Tender_manubriosternal_neg | Tender_manubriosternal_pos |
| | Tender_acromioclaviculair_L_neg | Tender_acromioclaviculair_L_pos |
| | Tender_acromioclaviculair_R_neg | Tender_acromioclaviculair_R_pos |
| | Tender_ankle_L_neg | Tender_ankle_L_pos |
| | Tender_ankle_R_neg | Tender_ankle_R_pos |
| | Tender_cervical spine_neg | Tender_cervical spine_pos |
| | Tender_CMC_L_neg | Tender_CMC_L_pos |
| | Tender_CMC_R_neg | Tender_CMC_R_pos |
| | Tender_DIP_2_hand_L_neg | Tender_DIP_2_hand_L_pos |
| | Tender_DIP_2_foot_L_neg | Tender_DIP_2_foot_L_pos |
| | Tender_DIP_2_hand_R_neg | Tender_DIP_2_hand_R_pos |
| | Tender_DIP_2_foot_R_neg | Tender_DIP_2_foot_R_pos |
| | Tender_DIP_3_hand_L_neg | Tender_DIP_3_hand_L_pos |
| | Tender_DIP_3_foot_L_neg | Tender_DIP_3_foot_L_pos |
| | Tender_DIP_3_hand_R_neg | Tender_DIP_3_hand_R_pos |
| | Tender_DIP_3_foot_R_neg | Tender_DIP_3_foot_R_pos |
| | Tender_DIP_4_hand_L_neg | Tender_DIP_4_hand_L_pos |
| | Tender_DIP_4_foot_L_neg | Tender_DIP_4_foot_L_pos |
| | Tender_DIP_4_hand_R_neg | Tender_DIP_4_hand_R_pos |
| | Tender_DIP_4_foot_R_neg | Tender_DIP_4_foot_R_pos |
| | Tender_DIP_5_hand_L_neg | Tender_DIP_5_hand_L_pos |
| | Tender_DIP_5_foot_L_neg | Tender_DIP_5_foot_L_pos |
| | Tender_DIP_5_hand_R_neg | Tender_DIP_5_hand_R_pos |
| | Tender_DIP_5_foot_R_neg | Tender_DIP_5_foot_R_pos |
| | Tender_elbow_R_neg | Tender_elbow_R_pos |
| | Tender_hip_L_neg | Tender_hip_L_pos |
| | Tender_hip_R_neg | Tender_hip_R_pos |
| | Tender_knee_L_neg | Tender_knee_L_pos |
| | Tender_knee_R_neg | Tender_knee_R_pos |
| | Tender_MCP_1_L_neg | Tender_MCP_1_L_pos |
| | Tender_MCP_1_R_neg | Tender_MCP_1_R_pos |
| | Tender_MCP_2_L_neg | Tender_MCP_2_L_pos |
| | Tender_MCP_2_R_neg | Tender_MCP_2_R_pos |
| | Tender_MCP_3_L_neg | Tender_MCP_3_L_pos |
| | Tender_MCP_3_R_neg | Tender_MCP_3_R_pos |
| | Tender_MCP_4_L_neg | Tender_MCP_4_L_pos |
| | Tender_MCP_4_R_neg | Tender_MCP_4_R_pos |
| | Tender_MCP_5_L_neg | Tender_MCP_5_L_pos |
| | Tender_MCP_5_R_neg | Tender_MCP_5_R_pos |
| | Tender_MTP_1_L_neg | Tender_MTP_1_L_pos |

| | | |
|---|---|---|
| | Tender_MTP_1_R_neg | Tender_MTP_1_R_pos |
| | Tender_MTP_2_L_neg | Tender_MTP_2_L_pos |
| | Tender_MTP_2_R_neg | Tender_MTP_2_R_pos |
| | Tender_MTP_3_L_neg | Tender_MTP_3_L_pos |
| | Tender_MTP_3_R_neg | Tender_MTP_3_R_pos |
| | Tender_MTP_4_L_neg | Tender_MTP_4_L_pos |
| | Tender_MTP_4_R_neg | Tender_MTP_4_R_pos |
| | Tender_MTP_5_L_neg | Tender_MTP_5_L_pos |
| | Tender_MTP_5_R_neg | Tender_MTP_5_R_pos |
| | Tender_talo-calcaneo-navicularis_L_neg | Tender_talo-calcaneo-navicularis_L_pos |
| | Tender_talo-calcaneo-navicularis_R_neg | Tender_talo-calcaneo-navicularis_R_pos |
| | Tender_PIP_2_hand_L_neg | Tender_PIP_2_hand_L_pos |
| | Tender_PIP_2_foot_L_neg | Tender_PIP_2_foot_L_pos |
| | Tender_PIP_2_hand_R_neg | Tender_PIP_2_hand_R_pos |
| | Tender_PIP_2_foot_R_neg | Tender_PIP_2_foot_R_pos |
| | Tender_PIP_3_hand_L_neg | Tender_PIP_3_hand_L_pos |
| | Tender_PIP_3_foot_L_neg | Tender_PIP_3_foot_L_pos |
| | Tender_PIP_3_hand_R_neg | Tender_PIP_3_hand_R_pos |
| | Tender_PIP_3_foot_R_neg | Tender_PIP_3_foot_R_pos |
| | Tender_PIP_4_hand_L_neg | Tender_PIP_4_hand_L_pos |
| | Tender_PIP_4_foot_L_neg | Tender_PIP_4_foot_L_pos |
| | Tender_PIP_4_hand_R_neg | Tender_PIP_4_hand_R_pos |
| | Tender_PIP_4_foot_R_neg | Tender_PIP_4_foot_R_pos |
| | Tender_PIP_5_hand_L_neg | Tender_PIP_5_hand_L_pos |
| | Tender_PIP_5_foot_L_neg | Tender_PIP_5_foot_L_pos |
| | Tender_PIP_5_hand_R_neg | Tender_PIP_5_hand_R_pos |
| | Tender_PIP_5_foot_R_neg | Tender_PIP_5_foot_R_pos |
| | Tender_wrist_L_neg | Tender_wrist_L_pos |
| | Tender_wrist_R_neg | Tender_wrist_R_pos |
| | Tender_sacroiliac_L_neg | Tender_sacroiliac_L_pos |
| | Tender_sacroiliac_R_neg | Tender_sacroiliac_R_pos |
| | Tender_shoulder_L_neg | Tender_shoulder_L_pos |
| | Tender_shoulder_R_neg | Tender_shoulder_R_pos |
| | Tender_sternoclavicular_L_neg | Tender_sternoclavicular_L_pos |
| | Tender_sternoclavicular_R_neg | Tender_sternoclavicular_R_pos |
| | Tender_tarsometatarsal_L_neg | Tender_tarsometatarsal_L_pos |
| | Tender_tarsometatarsal_R_neg | Tender_tarsometatarsal_R_pos |
| | Tender_temporomandibular_L_neg | Tender_temporomandibular_L_pos |
| | Tender_temporomandibular_R_neg | Tender_temporomandibular_R_pos |
| | Swollen_elbow_L_neg | Swollen_elbow_L_pos |
| | Swollen_IP_hand_L_neg | Swollen_IP_hand_L_pos |
| | Swollen_IP_hand_R_neg | Swollen_IP_hand_R_pos |
| | Swollen_IP_foot_L_neg | Swollen_IP_foot_L_pos |
| | Swollen_IP_foot_R_neg | Swollen_IP_foot_R_pos |
| | Swollen_acromioclaviculair_L_neg | Swollen_acromioclaviculair_L_pos |
| | Swollen_acromioclaviculair_R_neg | Swollen_acromioclaviculair_R_pos |

| | | |
|---|---|---|
| | Swollen_ankle_L_neg | Swollen_ankle_L_pos |
| **SEROLOGY** | ACPA_neg | ACPA_pos |
| | RF_neg | RF_pos |
| **LAB** | MCV (Mean Corposcular Volume)** | MCH (Mean Corposcular Hemoglobin)* |
| | MCHC (Mean Corposcular Hemoglobin Concentration)* | ESR (Erytrocyte Sedimentation Rate) |
| | Hemoglobin | Hematocrit* |
| | Leukocytes | Trombocytes |
| **DEMOGRAPHICS** | Age | Sex |

*Where L, left; R, right; neg, negative; pos, positive; IP, interphalangeal; CMC, carpometacarpal; DIP, distal interphalangeal; PIP, proximal Interphalangeal joints; MCP, metacarpophalangeal; MTP, metatarsophalangeal joints; RF, rheumatoid factor; ACPA, anti-cyclic citrullinated peptide (ACPA) antibodies; MCV, mean corpuscular volume; MCH, mean corpuscular hemoglobin; MCHC, mean corpuscular hemoglobin concentration; ESR, erythrocyte sedimentation rate;*

***Table S2. Baseline characteristics of the different patient clusters in set A.*** *The clinical variables used for clustering are marked with the gamma symbol (ɣ).*

| | C1-Foot | C2-Oligo | C3-Hand | C4-Poly |
|---|---|---|---|---|
| **N** | 415 | 380 | 323 | 269 |
| **Sex, female [ɣ] [N(%)]** | 262 (63.1) | 262 (68.9) | 198 (61.3) | 172 (63.9) |
| **Age [ɣ] (SD, yr)** | 56.8 (14.4) | 59.7 (14.9) | 68.5 (12.6) | 55.1 (14.7) |
| **RF [ɣ] [N(%)]** | 245 (59.0) | 236 (62.1) | 121 (37.5) | 120 (44.6) |
| **ACPA [ɣ] [N(%)]** | 241 (58.1) | 224 (58.9) | 97 (30.0) | 114 (42.4) |
| **ESR [ɣ] (IQR, mm/hr)** | 22 (9-36) | 29 (14-46) | 31 (17-51) | 22 (9-41) |
| **DAS44(3) (IQR)** | 3.5 (3.0-4.0) | 2.5 (2.0-2.8) | 3.8 (3.2-4.4) | 4.7 (4.0-5.5) |
| **SJC (IQR)** | 8 (5-11) | 3 (1-4) | 10 (7-14) | 14 (9-21) |
| **TJC (IQR)** | 11 (8-15) | 3 (2-5) | 11 (8-15) | 24 (18-31) |
| **DAS28(3) (IQR)** | 5.3 (4.4-6.0) | 4.2 (3.4-4.7) | 5.6 (5.0-6.4) | 6.6 (5.5-7.4) |
| **MTX [N(%)]** | 354 (85.3) | 275 (72.4) | 240 (74.3) | 218 (81.0) |
| **Follow up (IQR, days)** | 1749 (860-2663) | 1822 (1024-2587) | 1267 (706-2179) | 2032 (1141-2909) |
| **Symptom duration (IQR, days)** | 154 (56-365) | 217 (63-740) | 122 (42-365) | 155 (55-365) |

*SD= standard deviation; RF= Rheumatoid factor; ACPA= Anti-cyclic citrullinated peptide (ACPA) antibodies; ESR=erythrocyte sedimentation rate; IQR=interquartile range; DAS=Three component disease activity score (either 44 or 28 joint scheme); SJC=swollen joint count; TJC=tender joint count; MTX= prevalence of patients receiving methotrexate at baseline;*

***Table S3: Cluster table for replication set B (IMPROVED trial data).*** *The clinical variables used for clustering are marked with the gamma symbol (ɣ).*

| | C1-Foot | C2-Oligo | C3-Hand | C4-Poly |
|---|---|---|---|---|
| **N** | 90 | 102 | 50 | 65 |
| **Sex, female ˠ [N(%)]** | 66 (73.3) | 67 (65.7) | 28 (56.0) | 50 (76.9) |
| **Age ˠ (SD, yr)** | 51.0 (13.4) | 53.9 (13.5) | 58.1 (12.9) | 51.3 (15.8) |
| **RF ˠ [N(%)]** | 68 (75.6) | 75 (73.5) | 36 (72.0) | 40 (61.5) |
| **ACPA ˠ [N(%)]** | 63 (70.0) | 71 (69.6) | 29 (58.0) | 38 (58.5) |
| **ESR ˠ (IQR, mm/hr)** | 28 (11-41) | 29 (16-40) | 32 (16-50) | 33 (17-50) |
| **DAS44(3) (IQR)** | 3.3 (2.9-3.7) | 2.8 (2.5-3.0) | 3.3 (3.0-4.0) | 4.3 (3.7-4.7) |
| **SJC (IQR)** | 8 (5-11) | 4 (2-6) | 9 (6-12) | 16 (10-22) |
| **TJC (IQR)** | 7 (6-10) | 5 (3-7) | 8 (6-10) | 12 (9-15) |
| **DAS28(3) (IQR)** | 5.0 (4.5-5.8) | 4.6 (4.1-5.0) | 5.2 (4.8-5.8) | 6.1 (5.4-6.7) |
| **Follow up (IQR, days)** | 2119 (1018-3914) | 2553 (1521-4744) | 1730 (990-3258) | 2177 (1389-4368) |
| **Symptom duration (IQR, days)** | 161 (80-281) | 135 (67-258) | 144 (70-252) | 129 (75-252) |

*Where SD, standard deviation; RF, rheumatoid factor; ACPA, anti-cyclic citrullinated peptide antibodies; ESR, erythrocyte sedimentation rate; IQR, interquartile range; DAS, three component disease activity score (either 44 or 28 joint scheme); SJC, swollen joint count; TJC, tender joint count;*

***Table S4: Cluster table for replication set C (Reumazorg Zuid West Nederland hospital data).*** *The clinical variables used for clustering are marked with the gamma symbol (ɣ).*

| | C1-Foot | C2-Oligo | C3-Hand | C4-Poly |
|---|---|---|---|---|
| ***N*** | 91 | 279 | 77 | 68 |
| ***Sex, female ɣ [n(%)]*** | 61 (67.0) | 176 (63.1) | 46 (59.7) | 50 (73.5) |
| ***Age ɣ (SD, yr)*** | 61.3 (14.4) | 61.2 (14.2) | 63.3 (12.9) | 56.4 (13.3) |
| ***RF ɣ [n(%)]*** | 57 (62.6) | 186 (66.7) | 43 (55.8) | 36 (52.9) |
| ***ACPA ɣ [n(%)]*** | 51 (56.0) | 154 (55.2) | 36 (46.8) | 31 (45.6) |
| ***ESR ɣ (IQR, mm/hr)*** | 16 (8-26) | 17 (8-27) | 13 (7-29) | 14 (5-30) |
| ***DAS44(3) (IQR)*** | 3.8 (3.2-4.6) | 2.2 (1.8-2.6) | 3.7 (3.2-4.4) | 4.9 (4.2-5.9) |
| ***SJC (IQR)*** | 10 (6-15) | 2 (0-4) | 14 (10-17) | 18 (14-26) |
| ***TJC (IQR)*** | 14 (10-18) | 3 (1-5) | 11 (7-18) | 24 (17-38) |
| ***DAS28(3) (IQR)*** | 5.3 (4.6-6.3) | 3.6 (3.0-4.1) | 5.3 (4.7-6.0) | 6.3 (5.5-7.3) |
| ***Follow up (IQR, days)*** | 1382 (1047-1932) | 1515 (968-2183) | 1579 (1149-2275) | 1843 (1494-2256) |
| ***Symptom duration (IQR, days)*** | - | - | - | - |

***Table S5: Summary of Cox regression model based on potential mediators for association between clusters and MTX-failure after 1 year.***

| | Coef | Exp(Coef) | SE(Coef) | Z | P-value |
|---|---|---|---|---|---|
| ***SJC*** | -0.002 | 0.998 | 0.017 | -0.105 | 0.916 |
| ***TJC*** | -0.035 | 0.966 | 0.014 | -2.453 | 0.014 * |

| | | | | | |
|---|---|---|---|---|---|
| ***ACPA*** | 0.164 | 1.178 | 0.229 | 0.713 | 0.476 |
| ***RF*** | -0.071 | 0.932 | 0.222 | -0.318 | 0.751 |
| ***Sex, Female*** | 0.202 | 1.224 | 0.180 | 0.952 | 0.341 |
| ***ESR*** | 0.006 | 1.006 | 0.003 | 1.886 | 0.059 |
| ***Symptom duration*** | <0.001 | 1.001 | <0.001 | 2.256 | 0.024 * |
| ***Treatment delay*** | <0.001 | 0.998 | <0.001 | -0.406 | 0.684 |
| ***C1-Feet*** | -0.545 | 0.580 | 0.248 | -2.202 | 0.028 * |
| ***C2-Oligo*** | -1.232 | 0.292 | 0.354 | -3.482 | <0.001 * |
| ***C3-Hand*** | -1.366 | 0.255 | 0.312 | -4.379 | <0.001 * |
| ***C4-Poly*** | NA | NA | NA | NA | NA |

*Where Coef, regression coefficient; Exp(Coef), exponential of regression coefficient (also known as hazard ratio); SE, standard deviation; SJC, swollen joint count; TJC, tender joint count; ACPA, anti-cyclic citrullinated peptide antibodies; RF, Rheumatoid factor; ESR, erythrocyte sedimentation rate; NA, not available;*

***Table S6. Baseline characteristics of the final population compared to the dropped patients in set A (hospital data from Leiden University Medical Center)***

| | **Set A** | **Dropped patients** |
|---|---|---|
| **N** | 1387 | 1304 |
| **# Patients** | 1343 | 1082 |
| **Sex, Female (n(%))** | 894 (64.5) | 888 (68.3) |
| **Age (SD)** | 60.0 (15.0) | 59.9 (16.6) |
| **RF+ (n(%))** | 722 (52.1) | 453 (48.7) |
| **Missing RF (n(%))** | 0 (0.0) | 374 (28.7) |
| **ACPA+ (n(%))** | 676 (48.7) | 443 (46.4) |
| **Missing ACPA (n(%))** | 0 (0.0) | 350 (26.8) |
| **ESR (IQR)** | 25 (11-45) | 22 (9-41) |
| **Missing ESR (n(%))** | 0.0 (0.0) | 662 (50.8) |
| **SJC (IQR)** | 7 (4-12) | 6 (3-11) |
| **No Swollen (n(%))** | 72 (5.2) | 67 (10.1) |
| **Missing SJC (n(%))** | 0 (0.0) | 643 (49.3) |
| **TJC (IQR)** | 10 (5-17) | 9 (4-15) |
| **No Tender (n(%))** | 43 (3.1) | 41 (6.2) |
| **Missing TJC (n(%))** | 0 (0.0) | 643 (49.3) |
| **DAS28(3) (IQR)** | 4.1 (3.2-4.9) | 3.4 (2.4-4.5) |
| **Missing DAS28(3) (n(%))** | 0.0 (0.0) | 318 (24.4) |
| **DAS44(3) (IQR)** | 2.6 (2.0-3.3) | 2.1 (1.3-2.9) |
| **Missing DAS44(3) (n(%))** | 0.0 (0.0) | 318 (24.4) |
| **Follow-Up (IQR, days)** | 1470 (622-2458) | 1219 (346-2388) |

*Where SD, standard deviation; RF, rheumatoid factor; ACPA, anti-cyclic citrullinated peptide antibodies; ESR, erythrocyte sedimentation rate; IQR, interquartile range; DAS, three component disease activity score (either 44 or 28 joint scheme); SJC, swollen joint count; TJC, tender joint count;*

*Table S7. Baseline characteristics of the final population compared to the dropped patients in set C (Reumazorg Zuid West Nederland)*

| | Set C | Dropped patients |
|---|---|---|
| **# Patients** | 515 | 712 |
| **Sex, Female (n(%))** | 333 (64.7) | 458 (63.9) |
| **Age (SD)** | 60.9 (14.1) | 62.4 (14.0) |
| **RF+ (n(%))** | 322 (62.5) | 206 (67.5) |
| **Missing RF (n(%))** | 0 (0.0) | 407 (57.0) |
| **ACPA+ (n(%))** | 272(52.8) | 157 (51.5) |
| **Missing ACPA (n(%))** | 0 (0.0) | 407 (57.0) |
| **ESR (IQR)** | 16 (8-27) | 15 (8-29) |
| **Missing ESR (n(%))** | 0.0 (0.0) | 103 (14.5) |
| **SJC (IQR)** | 5 (1-13) | 6 (2-13) |
| **Missing SJC (n(%))** | 0 (0.0) | 294 (41.3) |
| **TJC (IQR)** | 6 (2-14.0) | 6 (2-15) |
| **Missing TJC (n(%))** | 0 (0.0) | 294 (41.3) |
| **DAS28(3) (IQR)** | 4.3 (3.4-5.5) | 4.4 (3.4-5.2) |
| **Missing DAS28(3) (n(%))** | 0.0 (0.0) | 377 (52.9) |
| **DAS44(3) (IQR)** | 2.8 (1.9-3.9) | 2.9 (2.1-3.8) |
| **Missing DAS44(3) (n(%))** | 0.0 (0.0) | 377 (52.9) |
| **Follow-Up (IQR, days)** | 1568 (1012-2174) | 1784 (823-2682) |

*Where SD, standard deviation; RF, rheumatoid factor; ACPA, anti-cyclic citrullinated peptide antibodies; ESR, erythrocyte sedimentation rate; IQR, interquartile range; DAS, three component disease activity score (either 44 or 28 joint scheme); SJC, swollen joint count; TJC, tender joint count;*
