## supplemental figures for "Location of joint involvement differentiates rheumatoid arthritis into different clinical subsets"

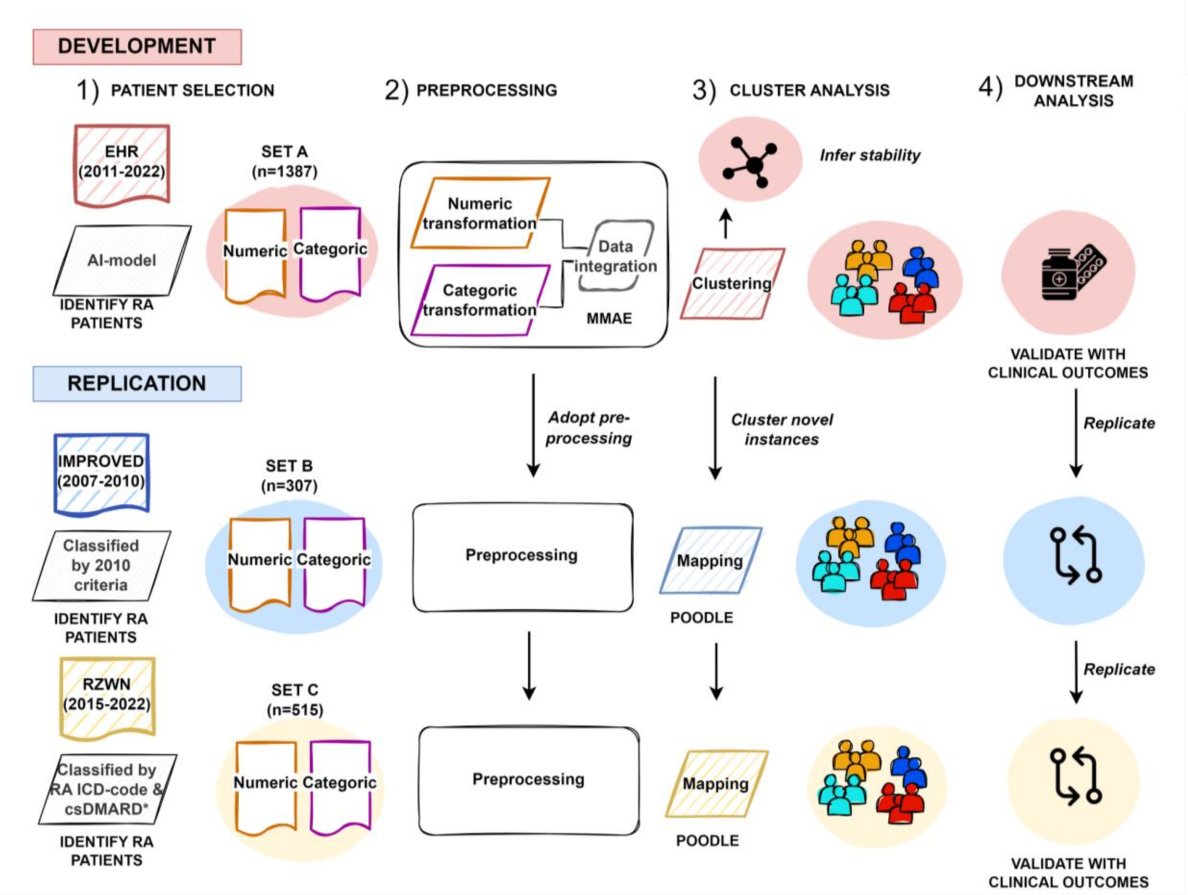


***Supplementary Fig. S1: Workflow depicting the different steps of our pipeline.*** *Where EHR, Electronic health records repository of Leiden University Medical Center (LUMC); AI, Artificial Intelligence, RZWN, Electronic health records repository of Reumazorg Zuid West Nederland; ICD, International Statistical Classification of Diseases; csDMARD, conventional Disease-Modifying Antirheumatic Drug.*

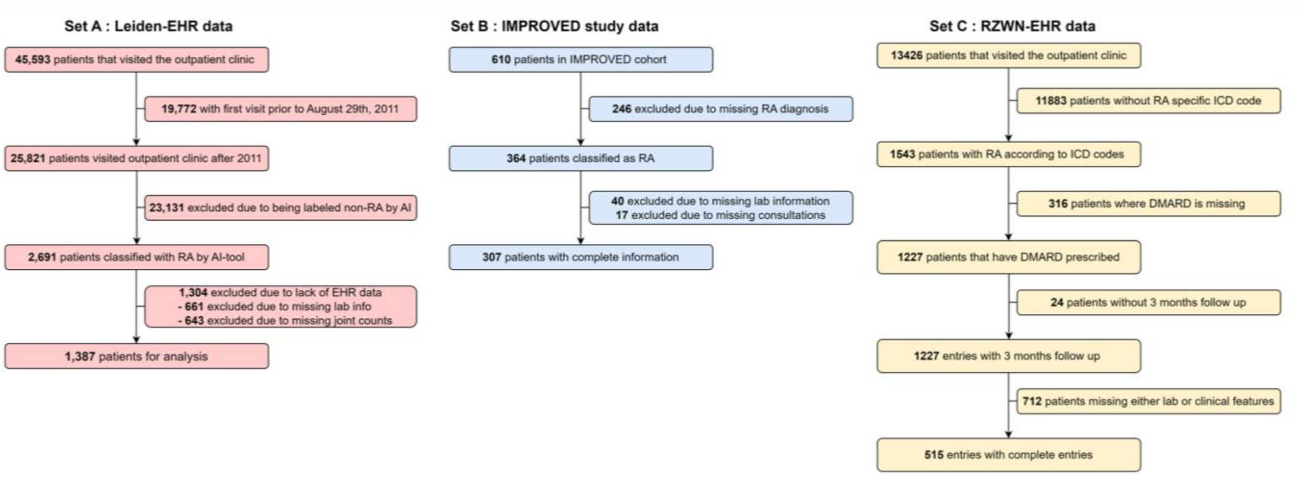


***Supplementary Fig. S2: Selection of patients for development (set A) and validation (set B & C), as formed from the electronic health records- and early arthritis clinic data respectively.*** *Where Leiden-EHR, Electronic health records repository of Leiden University Medical Center (LUMC); AI, Artificial Intelligence; RZWN-EHR, Electronic health records repository of Reumazorg Zuid West Nederland.*

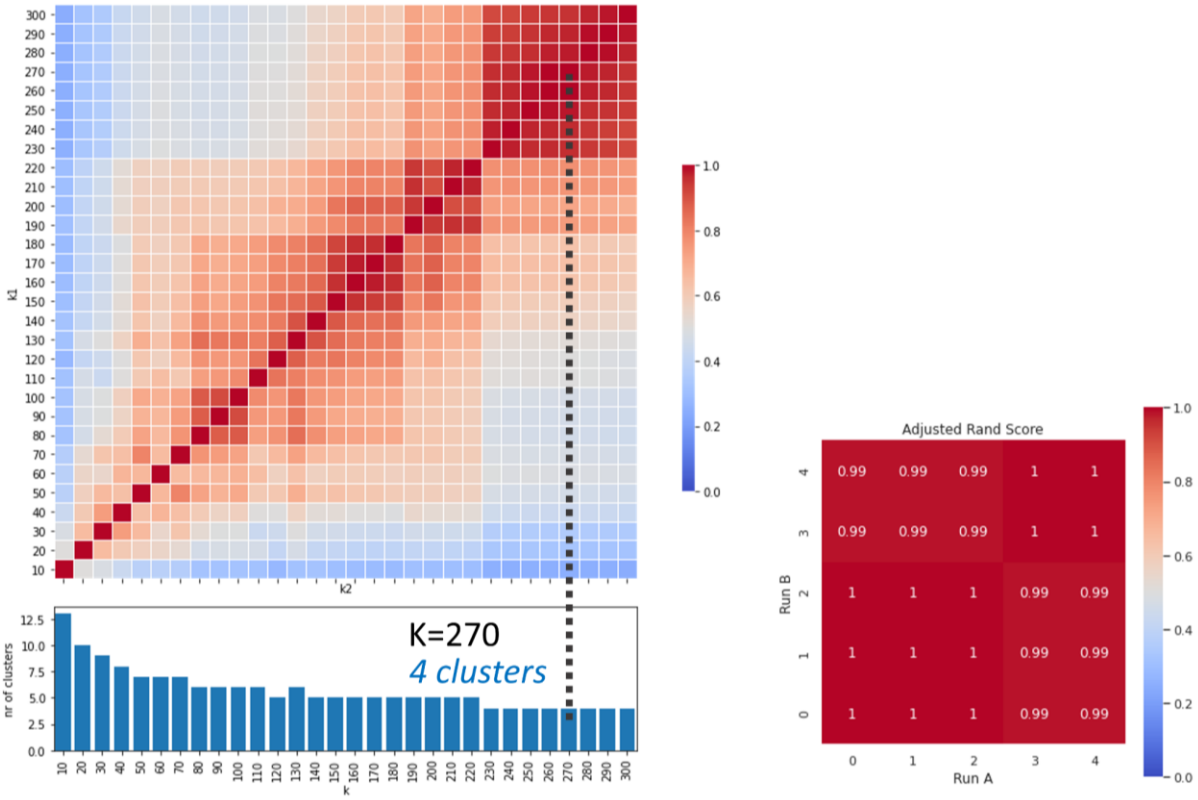


***Supplementary Fig. S3: Heatmap showing the similarity of clusterings across different configurations of k nearest neighbors (kNN), highlighting the stable and unstable regions for the k-parameter.*** *The similarity is expressed in adjusted rand index (ARI). The number of clusters associated with each configuration is displayed in the bar chart below. The dashed line indicates our choice of k and the corresponding number of clusters. Whereby k1 = k-value of the first setting of kNN and k2 = k-value of the second setting of kNN.*

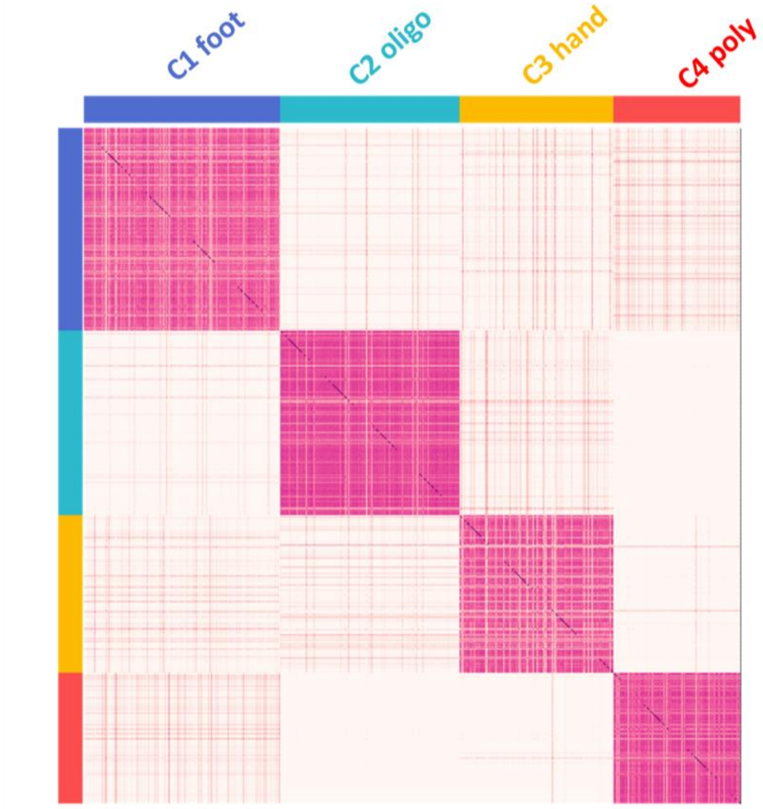


***Supplementary Fig. S4: Consensus heatmap depicting the cluster stability across 1000 bootstrapped iterations on random subsets of the data.*** *Pink denotes that patients co-clustered consistently, and white denotes that patients did not co-cluster. Patients are sorted according to their PhenoGraph clusters.*

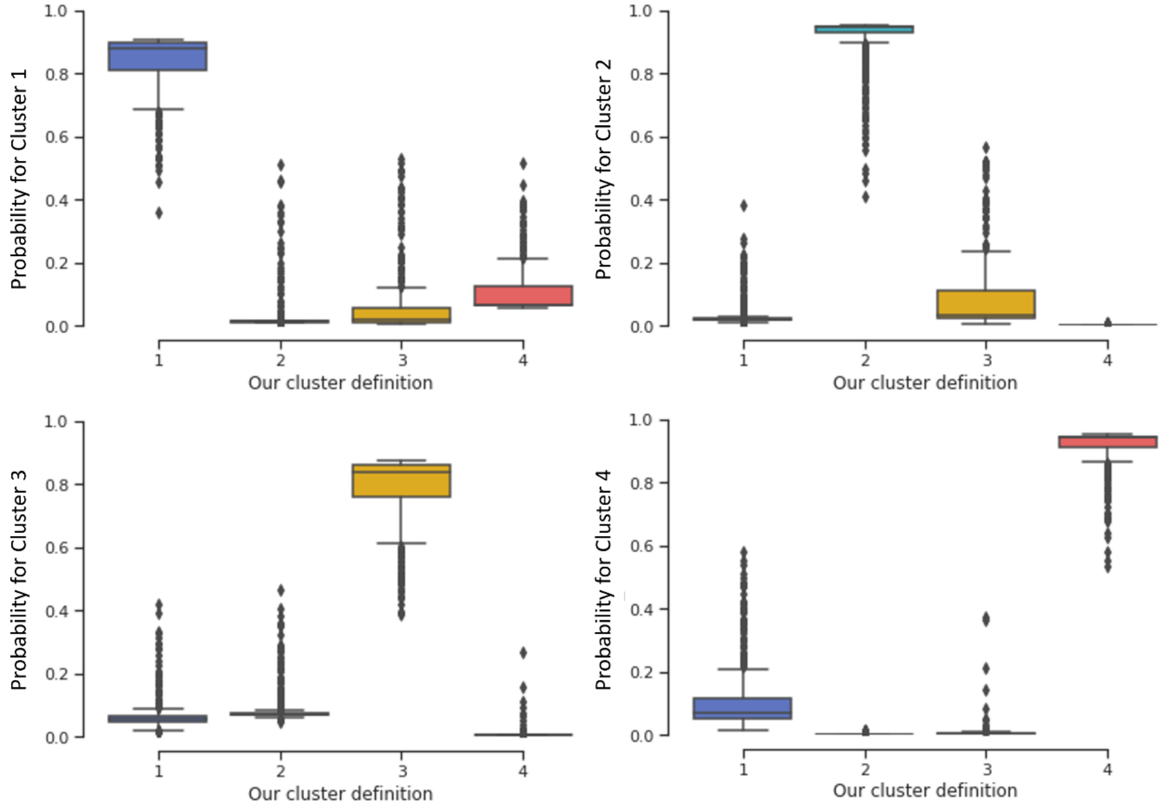


***Supplementary Fig. S5: Boxplots highlighting probability of patients ending up in the same subset of patients across the 1000 bootstrapping iterations.***

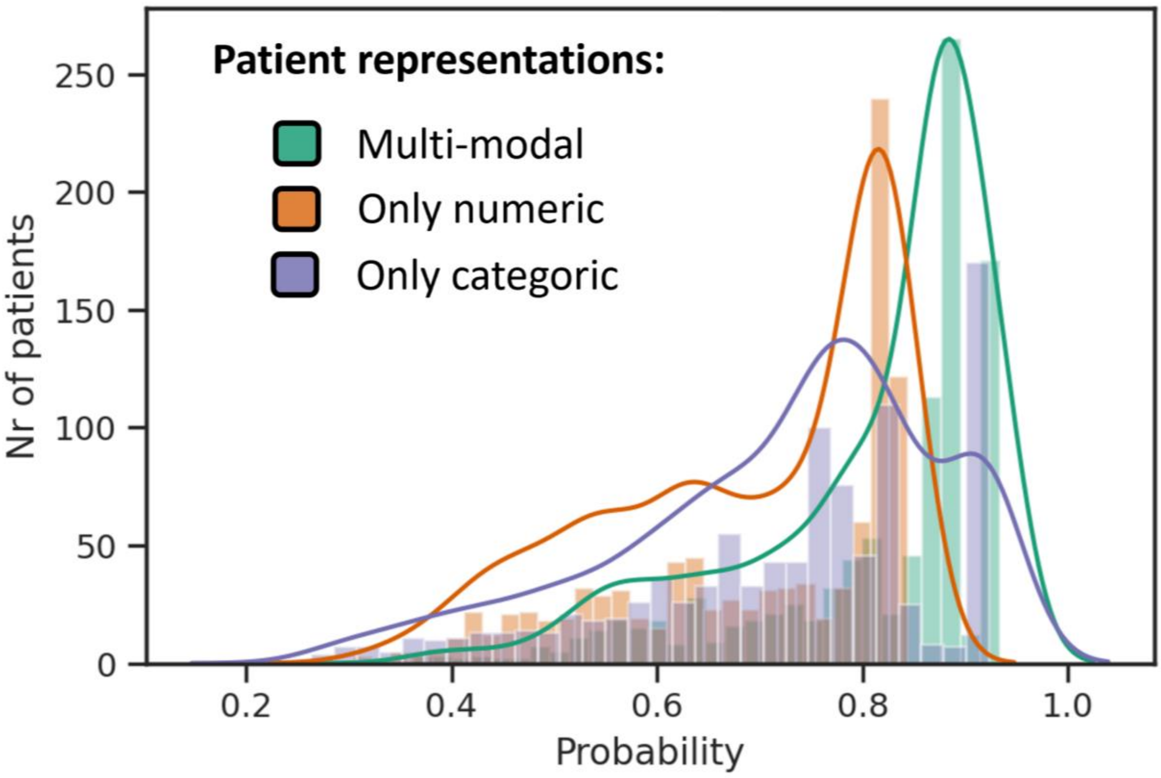

***Supplementary Fig. S6: Density plot highlighting probability of a patient ending up in the same cluster after bootstrapping 1000 times.*** *By generating random projections of the data, we can infer the stability of the clusters. We evaluated the stability of clustering for the different data types: numerical (lab, age), categorical (clinical, sex and autoantibodies) and the multi-modal approach that integrates both data types in a single feature space.*

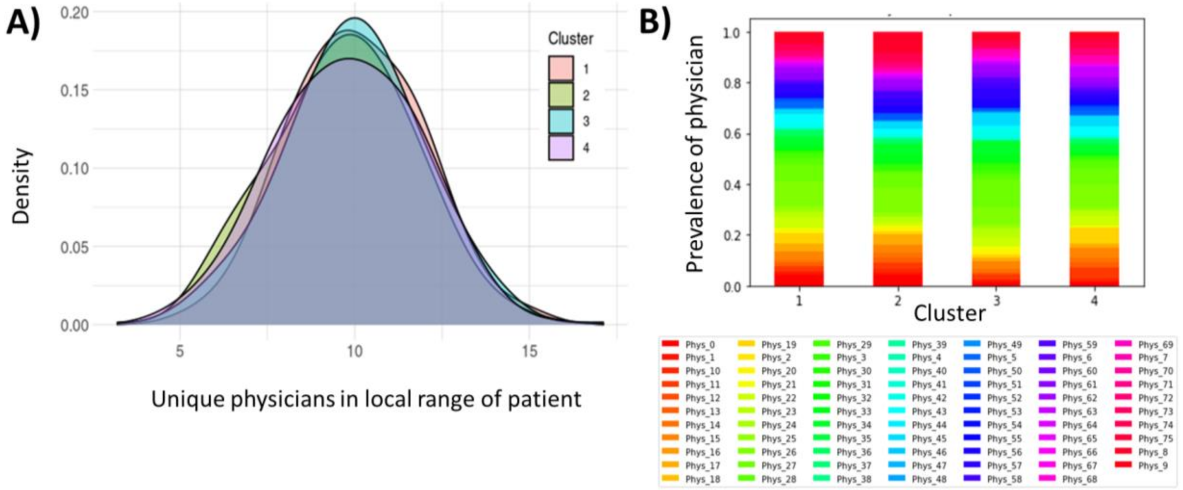


***Supplementary Fig. S7 Density plot showing the local diversity of physicians per cluster (A) and a stacked bar chart showing the prevalence of physician codes per cluster (B).*** *The local diversity is quantified with the Local Inverse Simpson's index (LISI), which shows the degree to which the physicians are well-mixed across neighboring data points (n=15 patients). We find that on average patients are surrounded by 10 unique physicians regardless of their cluster.*

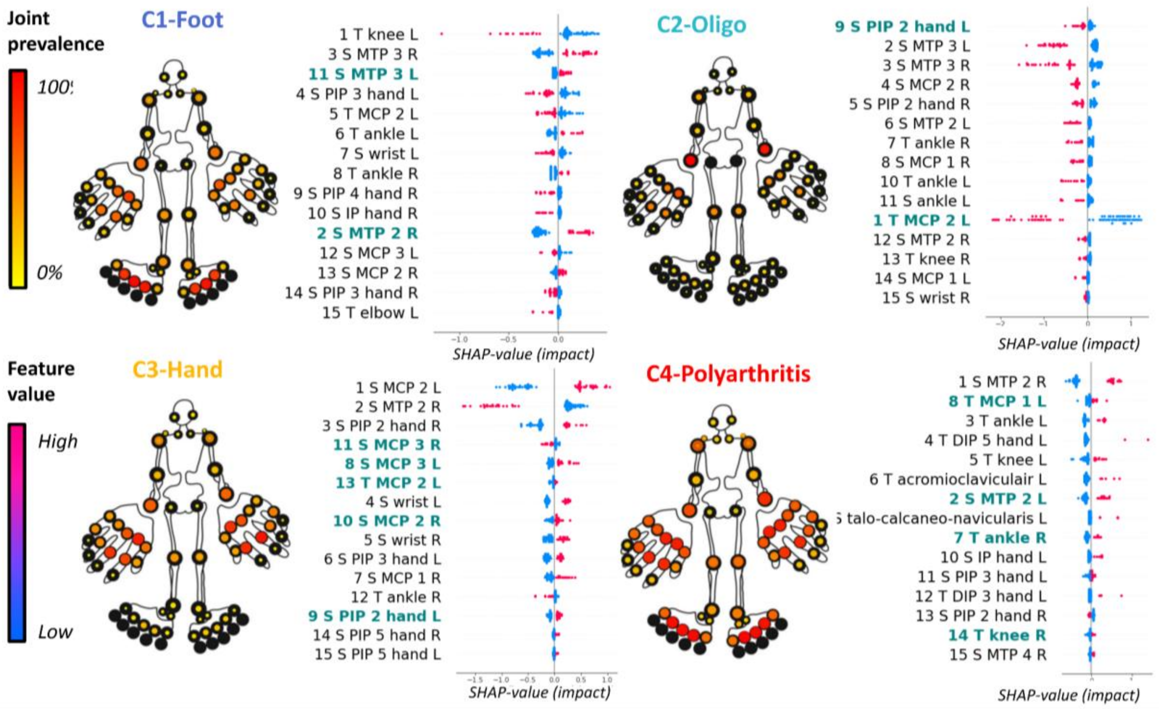

***Supplementary Fig S8: Cluster summary plots showing the average joint involvement and the top 15 driving variables in replication set B (IMPROVED).*** *The mannequin is formatted as a heatmap, showing the prevalence (red=100%, yellow=0%) of joint involvement (tender or swollen). In the SHAP plots, the most informative features for each cluster are listed in descending order. Here the x-axis shows the strength and direction of impact of that variable for each patient (represented by a dot), where initial value is represented with a color (pink=high, blue=low). The top 15 variables of the replication set are reordered and aligned, if possible, with the original set A (the prefix shows the rank in replication set). Variables that overlap are shown in bold, and colored on agreement (blue= same association, gold= inverse association). Where ACPA, anti-cyclic citrullinated peptide antibodies; ESR, erythrocyte sedimentation rate; IP, interphalangeal; L, left, MCH, mean corpuscular hemoglobin; MCHC, mean corpuscular hemoglobin concentration; MCP, metacarpophalangeal; MTP, metatarsophalangeal; PIP, proximal interphalangeal; R, right;*

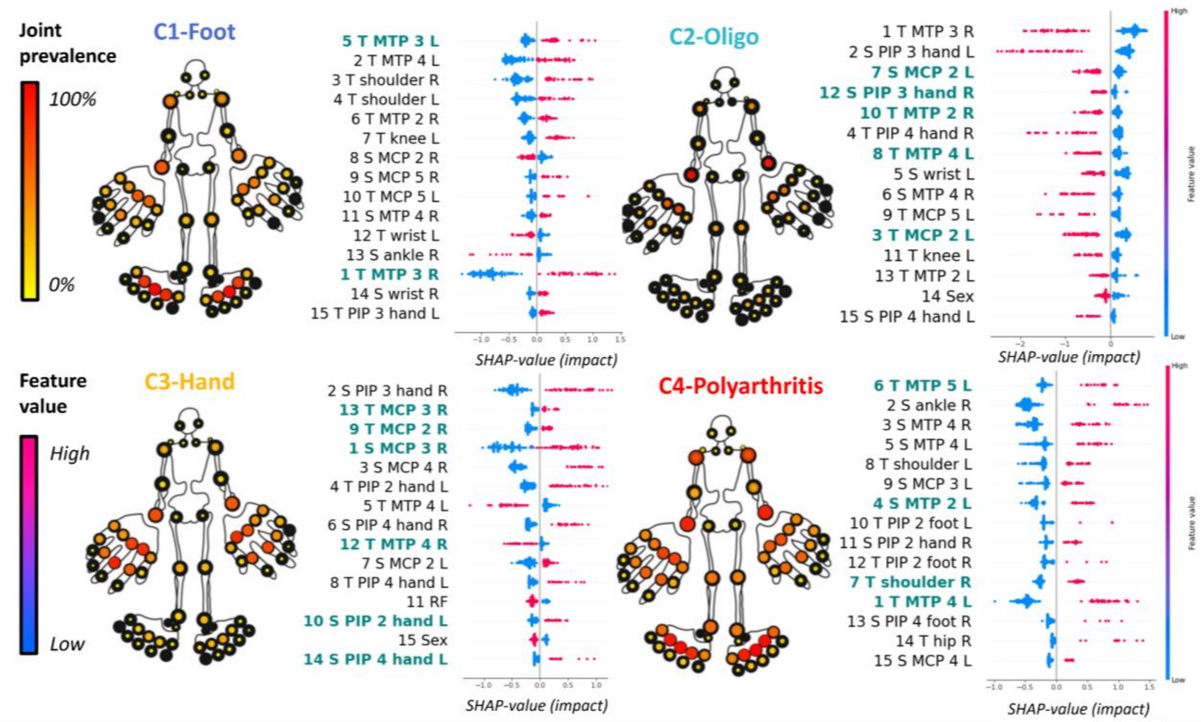


***Supplementary Fig S9: Cluster summary plots showing the average joint involvement and the top 15 driving variables in replication set C (RZWN-EHR).*** *The mannequin is formatted as a heatmap, showing the prevalence (red=100%, yellow=0%) of joint involvement (tender or swollen). In the SHAP plots, the most informative features for each cluster are listed in descending order. Here the x-axis shows the strength and direction of impact of that variable for each patient (represented by a dot), where initial value is represented with a color (pink=high, blue=low). The top 15 variables of the replication set are reordered and aligned, if possible, with the original set A (the prefix shows the rank in replication set). Variables that overlap are shown in bold, and colored on agreement (blue= same association, gold= inverse association). Where RF, rheumatoid factor; ESR, erythrocyte sedimentation rate; IP, interphalangeal; L, left; MCP, metacarpophalangeal; MTP, metatarsophalangeal; PIP, proximal interphalangeal; R, right;*

#### A) Set A – Leiden EHR (MTX-failure)

Model 1: Only baseline DAS44

| | coef | exp(coef) | se(coef) | z | P-Value |
|---|---|---|---|---|---|
| DAS44 | 0.146 | 1.157 | 0.090 | 1.617 | 0.106 |

Model 2: Cluster 3 & 4 + DAS44 at baseline

| | coef | exp(coef) | se(coef) | z | P-Value |
|---|---|---|---|---|---|
| DAS44 | -0.002 | 0.998 | 0.103 | -0.022 | 0.982 |
| CLUSTER 4 | 0.746 | 2.109 | 0.241 | 3.097 | 0.002* |

*P-value: 0.002* (ANOVA)*

| COX-regression analysis | |
|---|---|
| Dependent variable | event_1y (Time to MTX-failure within 1 year) |
| N | 438 (only MTX-starters from Cluster 3 & 4) |
| Number of events | 95 |

#### B) Set A – Leiden EHR (Remission)

Model 1: Only baseline DAS44

| | coef | exp(coef) | se(coef) | z | P-Value |
|---|---|---|---|---|---|
| DAS44 | -0.257 | 0.773 | 0.079 | -3.26 | 0.001 |

Model 2: Cluster 3 & 4 + DAS44 at baseline

| | coef | exp(coef) | se(coef) | z | P-Value |
|---|---|---|---|---|---|
| DAS44 | -0.173 | 0.841 | 0.089 | -1.956 | 0.051 |
| CLUSTER 4 | -0.389 | 0.678 | 0.186 | -2.087 | 0.037* |

*P-value: 0.035* (ANOVA)*

| COX-regression analysis | |
|---|---|
| Dependent variable | event_1y_Rem (Time to Remission within 1 year) |
| N | 359 (patients from Cluster 3 & 4) |
| Number of events | 150 |

#### C) Set C – RZWN EHR (MTX-failure)

Model 1: Only baseline DAS44

| | coef | exp(coef) | se(coef) | z | P-Value |
|---|---|---|---|---|---|
| DAS44 | 0.548 | 1.730 | 0.152 | 3.598 | 0.001* |

Model 2: Cluster 3 & 4 + DAS44 at baseline

| | coef | exp(coef) | se(coef) | z | P-Value |
|---|---|---|---|---|---|
| DAS44 | 0.408 | 1.504 | 0.161 | 2.538 | 0.011* |
| CLUSTER 4 | 0.643 | 1.902 | 0.303 | 2.120 | 0.034* |

*P-value: 0.033* (ANOVA)*

| COX-regression analysis | |
|---|---|
| Dependent variable | event_1y (Time to MTX-failure within 1 year) |
| N | 109 (only MTX-starters from Cluster 3 & 4) |
| Number of events | 54 |

#### D) Set B – IMPROVED (Remission)

Model 1: Only baseline DAS44

| | coef | exp(coef) | se(coef) | z | P-Value |
|---|---|---|---|---|---|
| DAS44 | -0.260 | 0.771 | 0.157 | -1.66 | 0.098 |

Model 2: Cluster 3 & 4 + DAS44 at baseline

| | coef | exp(coef) | se(coef) | z | P-Value |
|---|---|---|---|---|---|
| DAS44 | 0.012 | 1.013 | 0.182 | 0.069 | 0.945 |
| CLUSTER 4 | -0.923 | 0.397 | 0.296 | -3.116 | 0.002 * |

*P-value: 0.002* (ANOVA)*

| COX-regression analysis | |
|---|---|
| Dependent variable | event_1y_Rem (Time to Remission within 1 year) |
| N | 112 (patients from Cluster 3 & 4) |
| Number of events | 62 |

***Supplementary Fig S10: Comparison of different Cox-regression models to illustrate additive value of clusters on top of baseline DAS44 for predicting difference in time till MTX failure or Remission of the hand cluster (C3) vs polyarthritis cluster (C4).*** *Where model 1 shows the variance explained by baseline DAS44 and model 2 the additive effect of clustering on top of DAS44. Model 1 and model 2 are compared with an ANOVA (Analysis of variance) based on chi-squared test.*

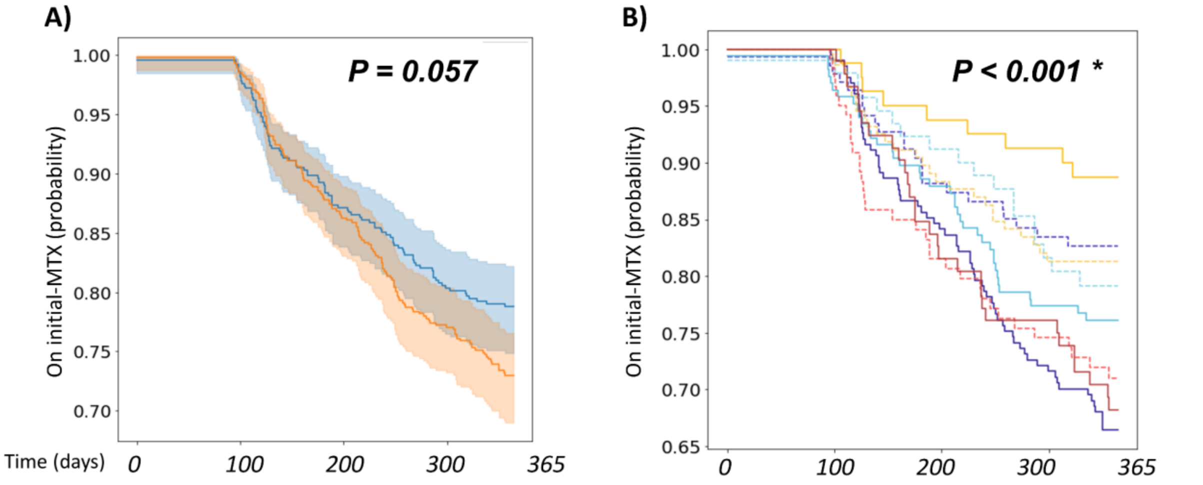


**NUMBER OF PATIENTS**

| Time (days) | 0 | 100 | 200 | 300 | 365 |
|---|---|---|---|---|---|
| ACPA- | 522 | 491 94% | 415 79% | 372 71% | 352 67% |
| ACPA+ | 562 | 534 95% | 460 82% | 403 72% | 374 66% |

**NUMBER OF PATIENTS**

| Time (days) | 0 | 100 | 200 | 300 | 365 |
|---|---|---|---|---|---|
| C1 ACPA+ | 211 | 201 95% | 169 80% | 140 66% | 129 61% |
| C1 ACPA- | 142 | 134 94% | 115 81% | 106 75% | 103 72% |
| C2 ACPA+ | 174 | 160 92% | 142 81% | 123 71% | 117 67% |
| C2 ACPA- | 99 | 91 92% | 80 81% | 67 68% | 63 64% |
| C3 ACPA+ | 84 | 81 96% | 74 88% | 72 86% | 69 82% |
| C3 ACPA- | 156 | 147 94% | 126 81% | 114 73% | 108 69% |
| C4 ACPA+ | 93 | 92 99% | 75 80% | 68 73% | 59 63% |
| C4 ACPA- | 125 | 119 95% | 94 75% | 85 68% | 78 62% |

***Supplementary Fig. S11: Kaplan Meier and survival data depicting global effect of ACPA on MTX switch rates (A), and local effect within clusters (B) in the Leiden EHR.*** *Here the dashed lines indicate the seropositive patients.*

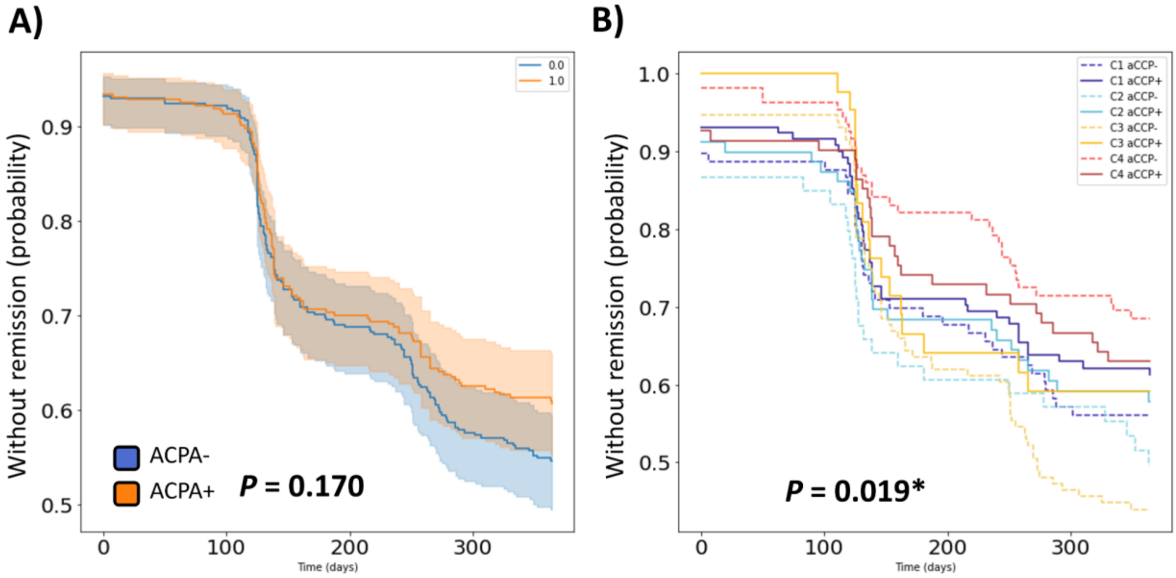


***Supplementary Fig S12: Kaplan Meier and survival data depicting global effect of ACPA on Remission rates (A), and local effect within clusters (B) in the Leiden EHR.*** *Here the dashed lines indicate the seropositive patients.*

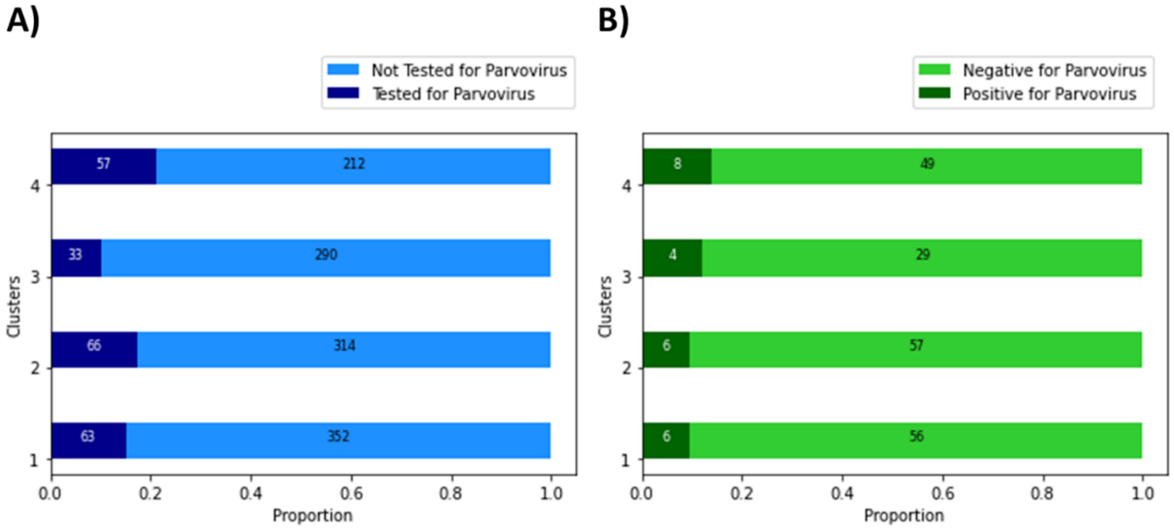


***Supplementary Fig. S13: Barchart depicting number of patients tested for parvovirus (A), and from those patients the number that tested positive (B)***

#### C3-hand vs C4-poly | C3-hand vs C1-Foot

### A) Set A – Leiden EHR

**C3-hand vs C4-poly**

Model 1: Only baseline DAS44

| | coef | exp(coef) | se(coef) | z | P-Value |
|---|---|---|---|---|---|
| DAS44 | 0.265 | 1.303 | 0.146 | 1.812 | 0.07 |

Model 2: Cluster 3 & 4 + DAS44 at baseline

| | coef | exp(coef) | se(coef) | Z | P-Value |
|---|---|---|---|---|---|
| DAS44 | 0.089 | 1.093 | 0.161 | 0.550 | 0.582 |
| CLUSTER 3 | -1.024 | 0.359 | 0.434 | -2.350 | 0.018* |

*P-value: 0.002* (ANOVA)*

| COX-regression analysis | |
|---|---|
| Dependent variable | event_1y (Time to MTX-failure within 1 year) |
| N | 167 (ACPA positive patients from Cluster 3 & 4) |
| Number of events | 36 |

**C3-hand vs C1-Foot**

Model 1: Only baseline DAS44

| | coef | exp(coef) | se(coef) | z | P-Value |
|---|---|---|---|---|---|
| DAS44 | -0.034 | 0.967 | 0.134 | -0.252 | 0.801 |

Model 2: Cluster 1 & 3 + DAS44 at baseline

| | coef | exp(coef) | se(coef) | Z | P-Value |
|---|---|---|---|---|---|
| DAS44 | 0.021 | 1.020 | 0.138 | 0.150 | 0.881 |
| CLUSTER 3 | -1.162 | 0.313 | 0.375 | -3.101 | 0.002* |

*P-value: >0.001 (ANOVA)*

| COX-regression analysis | |
|---|---|
| Dependent variable | event_1y (Time to MTX-failure within 1 year) |
| N | 306 (ACPA positive patients from Cluster 1 & 3) |
| Number of events | 78 |

### B) Set B – IMPROVED

**C3-hand vs C4-poly**

Model 1: Only baseline DAS44

| | coef (CI 95%) | std error | Z | P-Value |
|---|---|---|---|---|
| Intercept | -0.991 (-3.506, 1.523) | 1.283 | -0.773 | 0.440 |
| DAS44 | 0.216 (-0.433,0.864) | 0.331 | 0.651 | 0.515 |

Model 2: Cluster 3 & 4 + DAS44 at baseline

| | coef (CI 95%) | std error | Z | P-Value |
|---|---|---|---|---|
| Intercept | 1.863 (-1.517, 5.242) | 2.186 | 1.080 | 0.280 |
| CLUSTER 3 | -1.834 (-3.214, -0.455) | 0.704 | -2.606 | 0.009* |
| DAS44 | -0.345 (-0.448, -1.138) | 0.405 | -0.853 | 0.394 |

*P-value: 0.006* ($X^2$-test)*

| Logit regression analysis | |
|---|---|
| Dependent variable | event_1y_bin (MTX-failure within 1 year) |
| N | 64 (ACPA positive patients from Cluster 3 & 4) |
| Number of events | 26 |

**C3-hand vs C1-Foot**

Model 1: Only baseline DAS44

| | coef (CI 95%) | std error | Z | P-Value |
|---|---|---|---|---|
| Intercept | -1.425 (-4.167, 1.317) | 1.399 | -1.019 | 0.308 |
| DAS44 | 0.107 (-0.733, 0.947) | 0.429 | 0.250 | 0.803 |

Model 2: Cluster 1 & 3 + DAS44 at baseline

| | coef (CI 95%) | std error | Z | P-Value |
|---|---|---|---|---|
| Intercept | -1.425 (-4.168, 1.318) | 1.399 | -1.018 | 0.309 |
| CLUSTER 3 | -0.039 (-1.141, 1.063) | 0.562 | -0.069 | 0.945 |
| DAS44 | 0.110 (-0.735, 0.956) | 0.432 | 0.256 | 0.798 |

*P-value: 0.945 ($X^2$-test)*

| Logit regression analysis | |
|---|---|
| Dependent variable | event_1y_bin (MTX-failure within 1 year) |
| N | 88 (ACPA positive patients from Cluster 1 & 3) |
| Number of events | 21 |

### C) Set C – RZWN EHR

**C3-hand vs C4-poly**

Model 1: Only baseline DAS44

| | coef | exp(coef) | se(coef) | z | P-Value |
|---|---|---|---|---|---|
| DAS44 | 0.603 | 1.828 | 0.295 | 2.044 | 0.041* |

Model 2: Cluster 3 & 4 + DAS44 at baseline

| | coef | exp(coef) | se(coef) | z | P-Value |
|---|---|---|---|---|---|
| DAS44 | 0.559 | 1.748 | 0.271 | -2.063 | 0.039 * |
| CLUSTER 3 | -1.100 | 0.333 | 0.491 | -2.240 | 0.025 * |

*P-value: 0.019* (ANOVA)*

| COX-regression analysis | |
|---|---|
| Dependent variable | event_1y (Time to MTX-failure within 1 year) |
| N | 52 (ACPA positive patients from Cluster 3 & 4) |
| Number of events | 20 |

**C3-hand vs C1-Foot**

Model 1: Only baseline DAS44

| | coef | exp(coef) | se(coef) | z | P-Value |
|---|---|---|---|---|---|
| DAS44 | -0.215 | 0.806 | 0.222 | -0.968 | 0.333 |

Model 2: Cluster 1 & 3 + DAS44 at baseline

| | coef | exp(coef) | se(coef) | z | P-Value |
|---|---|---|---|---|---|
| DAS44 | -0.077 | 0.926 | 0.231 | -0.335 | 0.738 |
| CLUSTER 3 | -0.945 | 0.389 | 0.477 | -1.980 | 0.048* |

*P-value: 0.035* (ANOVA)*

| COX-regression analysis | |
|---|---|
| Dependent variable | event_1y (Time to MTX-failure within 1 year) |
| N | 66 (ACPA positive patients from Cluster 1 & 3) |
| Number of events | 27 |

***Suppl Fig S14:** Comparison of different regression models to illustrate additive value of clusters on top of baseline DAS44 for predicting MTX failure within ACPA stratum in A) Leiden hospital data B) IMPROVED trial*

***data and C) Reumazorg Zuid West Nederland hospital data.*** *Where we model the MTX-failure according to the difference between Cluster-3 and Cluster-4 or Cluster-1 with a Cox-regression (set A and C) or Logit regression technique (set B) in case of a fixed time of treatment switch. Model 1 shows the variance explained by baseline DAS44 and model 2 the additive effect of clustering on top of DAS44. Model 1 and model 2 are compared with an ANOVA (Analysis of variance) based on chi-squared test.*

Model 1: Only ACPA

| | coef | exp(coef) | se(coef) | z | P-Value |
|---|---|---|---|---|---|
| **ACPA** | 0.245 | 1.278 | 0.129 | 1.898 | 0.058 |

Model 2: ACPA adjusted for all other covariates

| | coef | exp(coef) | se(coef) | z | P-Value |
|---|---|---|---|---|---|
| **ACPA** | 0.376 | 1.456 | 0.172 | 2.190 | 0.029* |
| **RF** | -0.233 | 0.792 | 0.170 | -1.373 | 0.169 |
| **Age** | -0.010 | 0.989 | 0.004 | -2.331 | 0.020* |
| **Sex, Female** | 0.128 | 1.137 | 0.136 | 0.938 | 0.348 |

Model 3: Cluster + ACPA

| | coef | exp(coef) | se(coef) | z | P-Value |
|---|---|---|---|---|---|
| **C2-Oligo** | -0.203 | 0.816 | 0.170 | -1.195 | 0.232 |
| **C3-Hand** | -0.471 | 0.624 | 0.205 | -2.305 | 0.021* |
| **C4-Poly** | 0.173 | 1.189 | 0.165 | 1.045 | 0.296 |
| **ACPA** | 0.355 | 1.427 | 0.172 | 2.065 | 0.039* |
| **RF** | -0.221 | 0.802 | 0 171 | -1.291 | 0.197 |
| **Age** | -0.007 | 0.993 | 0.005 | -1.424 | 0.155 |
| **Sex, Female** | 0.159 | 1.172 | 0.137 | 1.155 | 0.248 |

***P-value** = 0.013* (ANOVA)*

| | Analysis of variance between Model 2 vs 3 |
|---|---|
| **Cox model** | *Response is Surv(time, event_1y)* |
| **Model 2** | *~ ACPA + RF + Age + Sex* |
| **Model 3** | *~ Cluster + ACPA + RF + Age + Sex* |
| **Statistical test** | ANOVA based on $\chi^2$ test |

| | COX-regression analysis |
|---|---|
| **Dependent variable** | *event_1y (Time to MTX-failure within 1 year)* |
| **N** | 1087 (only MTX-starters) |
| **Number of events** | 247 |

***Supplementary Fig. S15**: **Comparison of different Cox-regression models to illustrate additive value of clusters on top of ACPA for prediction of time till MTX failure.** Where model 1 shows the effect of ACPA-only, model 2 the effect of ACPA adjusted for other covariates, and model 3 the effect of clustering on top of covariates. Model 2 and model 3 are compared with an ANOVA (Analysis of variance) based on chi-squared test.*

**A)**

| | Coef | Exp(Coef) | Se(Coef) | Z | P-Value |
|---|---|---|---|---|---|
| **RF** | -0.236 | 0.790 | 0.171 | -1.379 | 0.168 |
| **ACPA** | 0.347 | 1.415 | 0.172 | 2.020 | 0.043* |
| **Age** | -0.008 | 0.992 | 0.005 | -1.776 | 0.076 |
| **Sex, F** | 0.150 | 1.161 | 0.140 | 1.066 | 0.287 |
| **SJC** | -0.004 | 0.996 | 0.013 | -0.339 | 0.734 |
| **TJC** | -0.004 | 0.995 | 0.011 | -0.418 | 0.676 |
| **BSE** | 0.006 | 1.006 | 0.002 | 2.602 | 0.009* |
| **C1-Foot** | -0.225 | 0.798 | 0.207 | -1.087 | 0.277 |
| **C2-Oligo** | -0.529 | 0.589 | 0.292 | -1.811 | 0.070 |
| **C3-Hand** | -0.750 | 0.472 | 0.263 | -2.854 | 0.004* |
| **C4-Poly** | NA | NA | NA | NA | NA |

1
2

**B)**

| | Coef | Exp(Coef) | Se(Coef) | Z | P-Value |
|---|---|---|---|---|---|
| **RF** | -0.229 | 0.795 | 0.170 | -1.347 | 0.178 |
| **ACPA** | 0.320 | 1.377 | 0.172 | 1.856 | 0.063 |
| **Age** | -0.007 | 0.992 | 0.005 | -1.640 | 0.101 |
| **Sex, F** | -0.111 | 1.118 | 0.142 | 0.783 | 0.434 |
| **SJC** | -0.003 | 0.997 | 0.022 | -0.150 | 0.881 |
| **TJC** | -0.002 | 0.998 | 0.020 | -0.091 | 0.927 |
| **BSE** | 0.006 | 1.006 | 0.002 | 2.546 | 0.011* |
| **Hand-JC** | -0.011 | 0.989 | 0.020 | -0.546 | 0.585 |
| **Feet-JC** | -0.019 | 1.019 | 0.021 | 0.923 | 0.356 |
| **C1-Foot** | -0.154 | 0.858 | 0.209 | -0.734 | 0.463 |
| **C2-Oligo** | -0.380 | 0.684 | 0.298 | -1.276 | 0.202 |
| **C3-Hand** | -0.501 | 0.606 | 0.298 | -1.682 | 0.093 |
| **C4-Poly** | NA | NA | NA | NA | NA |

3
4

**C)**

| | Model 1 | Model 2 | Model 3 |
|---|---|---|---|
| **Model 1** | | | |
| **Model 2** | 0.020 | | |
| **Model 3** | 0.008 | 0.637 | |
| **Model 4** | 0.023 | 0.202 | 0.331 |

***Supplementary Fig. S16: Comparison of Cox regression models for predicting MTX-failure after 1 year based on baseline predictors and cluster information (A) plus joint location counts in feet and hand (B).*** *To demonstrate the additive effects of our subsets, we compared our model 2 (including cluster information) to our baseline model, which constitutes the known predictors of MTX failure. Similarly, the hand/feet count model 3 was tested against complete model 4, to show the remaining effect of clustering after inclusion of joint location information. Statistical significance between models was inferred with ANOVA (C), where p<0.05 is marked with *. Where Coef, regression coefficient; Exp(Coef), exponential of regression coefficient (also known as hazard ratio); RF, rheumatoid factor; ACPA, anti-cyclic citrullinated peptide (ACPA) antibodies; ESR, erythrocyte sedimentation rate; SJC, swollen joint count; TJC, tender joint count; Hand-JC, number of affected hand joints; Feet-JC, number of affected feet joints; NA, not available;*

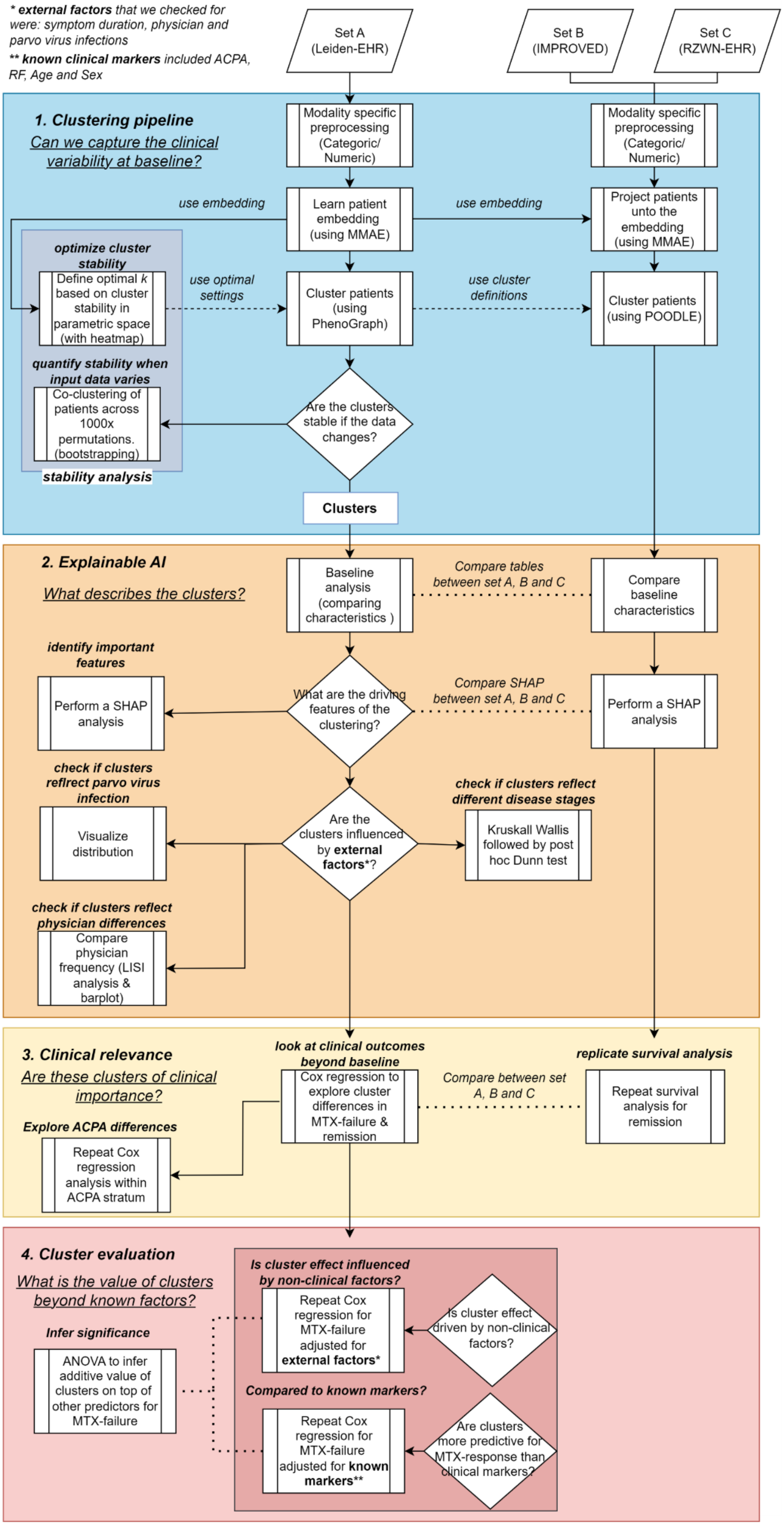


***Supplementary Fig. S17: Technical flow highlighting the important methodological steps of our study: 1) Creating clusters, 2) Explainable AI, 3) Clinical relevance, 4) Cluster evaluation.*** *Where EHR, Electronic health*

*records repository of Leiden University Medical Center (LUMC); AI, Artificial Intelligence; RZWN, hospital data from Reumazorg Zuid West Nederland;*

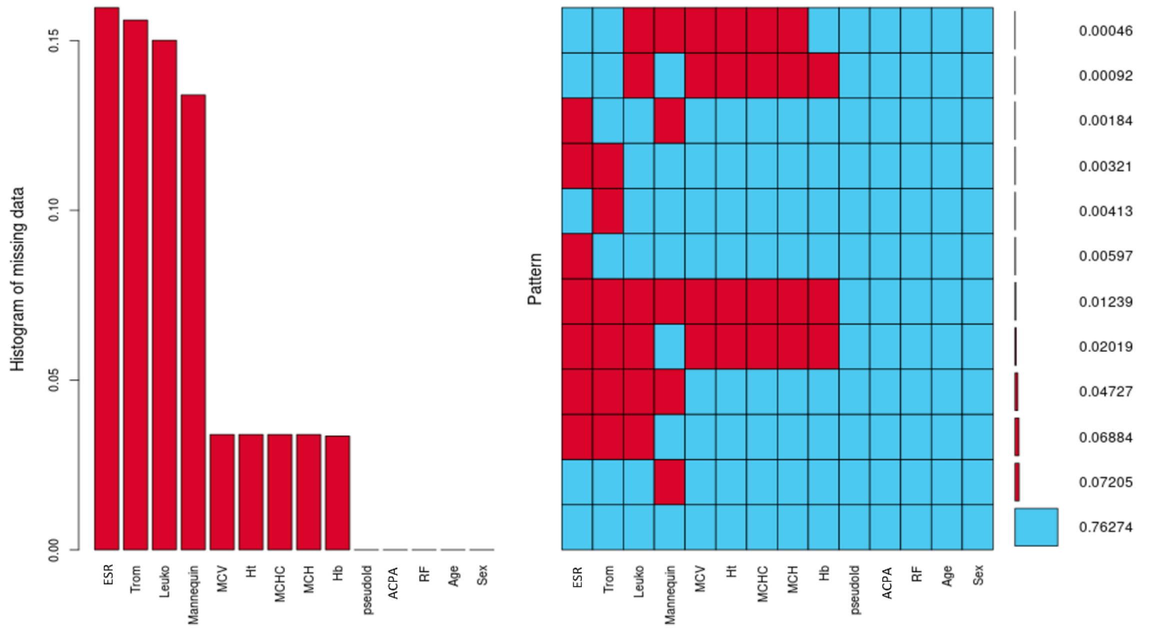


***Supplementary Fig. S18: Most frequent patterns of missingness in the clinical and lab variables of the electronic health records from Leiden.*** *The histogram shows the absolute sum of missingness for each variable, whereas the heatmap illustrates the most prevalent patterns of missingness across rows. Where ESR, erythrocyte sedimentation rate; Trom, thrombocytes; Leuko, leukocytes; MCV, mean corpuscular volume; Ht, hematocrit; MCHC, mean corpuscular hemoglobin concentration; MCH, mean corpuscular hemoglobin; Hb, hemoglobin; ACPA, anti-cyclic citrullinated peptide antibodies; RF, rheumatoid factor;*

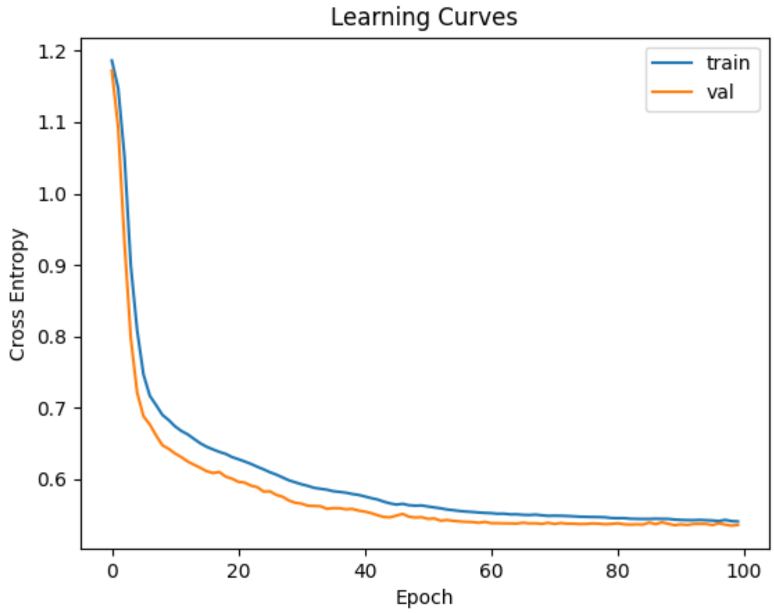


***Supplementary Fig. S19: Learning curves of the autoencoder depicting the reconstruction loss for both the training & validation set.***

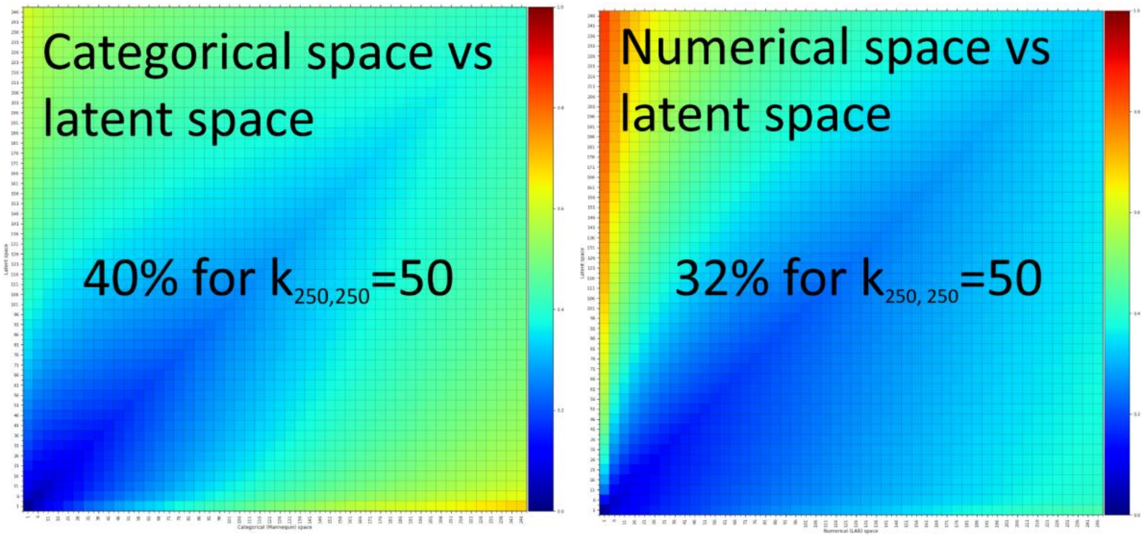


***Supplementary Fig. S20: Heatmap showing the conservation of local structure from the categorical- and numerical variable sets in the deep learned product space.*** *The preservation of local structure is measured with the scale dependent similarity measure across different configurations of the k-nearest neighbours algorithm (kNN).*

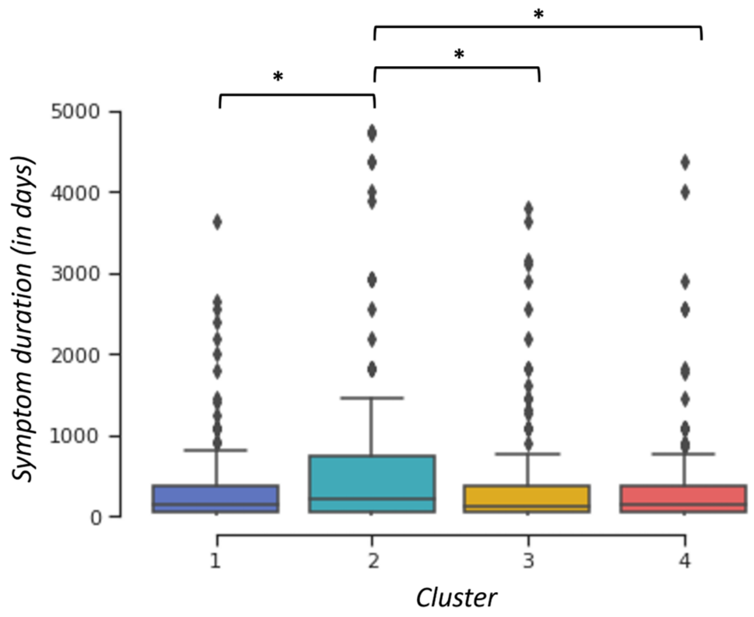


***Supplementary Fig. S21: Boxplots showing symptom duration till baseline across the different clusters.*** *Statistical significance was inferred with the Kruskall Wallis followed up by a Dunn test, where p<0.05 is marked with **

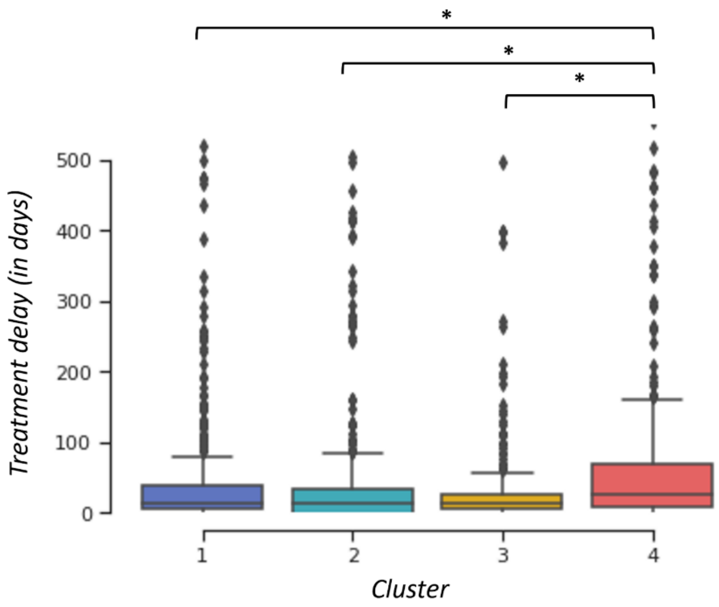


***Supplementary Fig. S22: Boxplot displaying the time from baseline till the start of the treatment.*** *Statistical significance was inferred with the Kruskall Wallis followed up by a Dunn test, where p<0.05 is marked with **

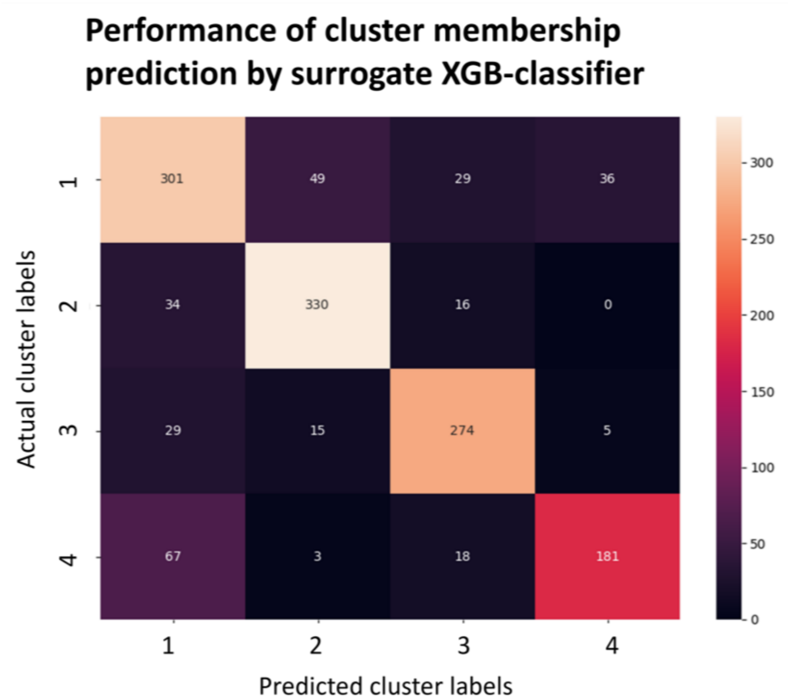


*Supplementary Fig. S23: Confusion matrix showing the ability of the surrogate XGB (eXtreme Gradient Boosting) classifier to predict the clusters and assess the SHAP values.*

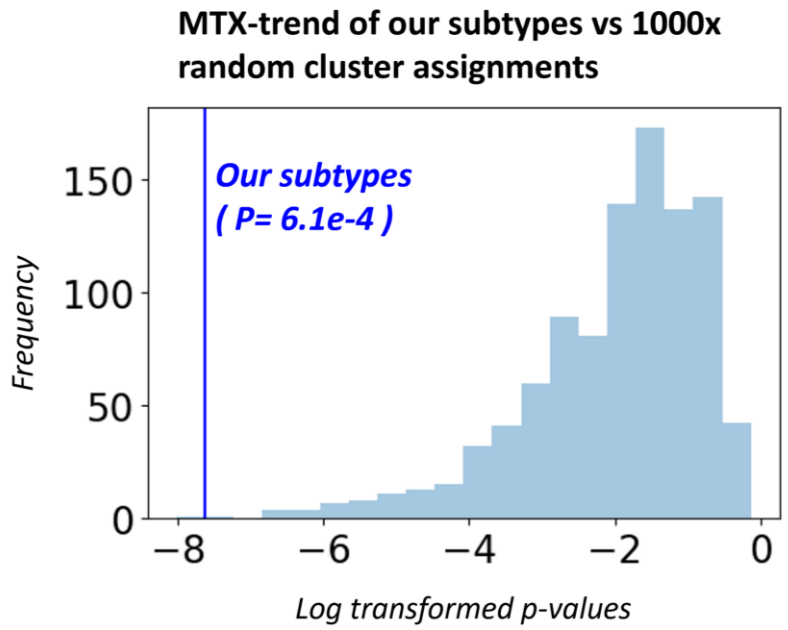


*Supplementary Fig. S24: Overrepresentation analysis where patients are randomly assigned to clusters 1000 times to see the likelihood of finding a similar Methotrexate-trend by random luck.*

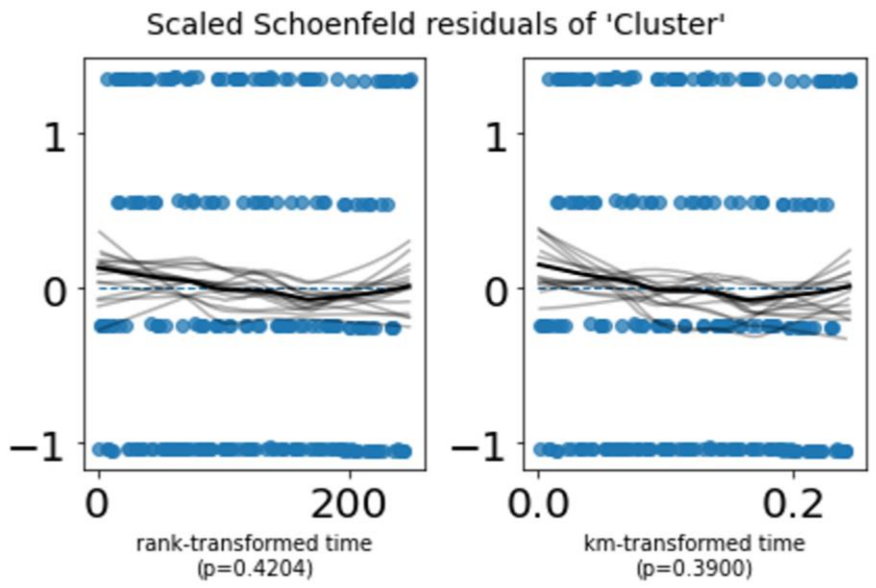


***Supplementary Fig. S25: Scaled Schoenfeld residual plots to examine whether or not the proportional hazard assumption holds.*** *We tested the residuals versus different functions of time for the clusters whereby a LOWESS (Locally Weighted Scatterplot Smoothing) line is fitted alongside 10 bootstrap LOWESS lines to check for non-zero slopes. Statistical significance was inferred with a chi-squared test. Here km=Kaplan Meier transformation; p=p-value; rank=log rank transformation.*

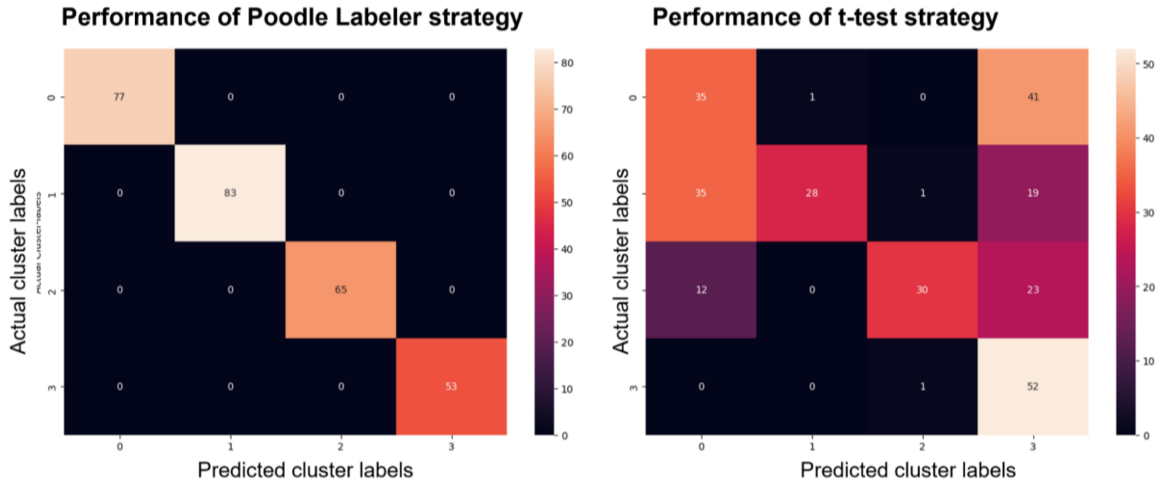


***Supplementary Fig. S26: Confusion matrix of POODLE labeler vs dummy strategy (assignment based on t-statistic) when predicting the cluster membership on a hold-out test set.*** *The POODLE technique assigns patients based on their similarity with the cluster, with respect to the average similarity between members of that cluster (within cluster similarity). In contrast, the dummy -test strategy applies a one-tailed unpaired t-test, and assigns patients to the cluster based on the largest t-statistic.*

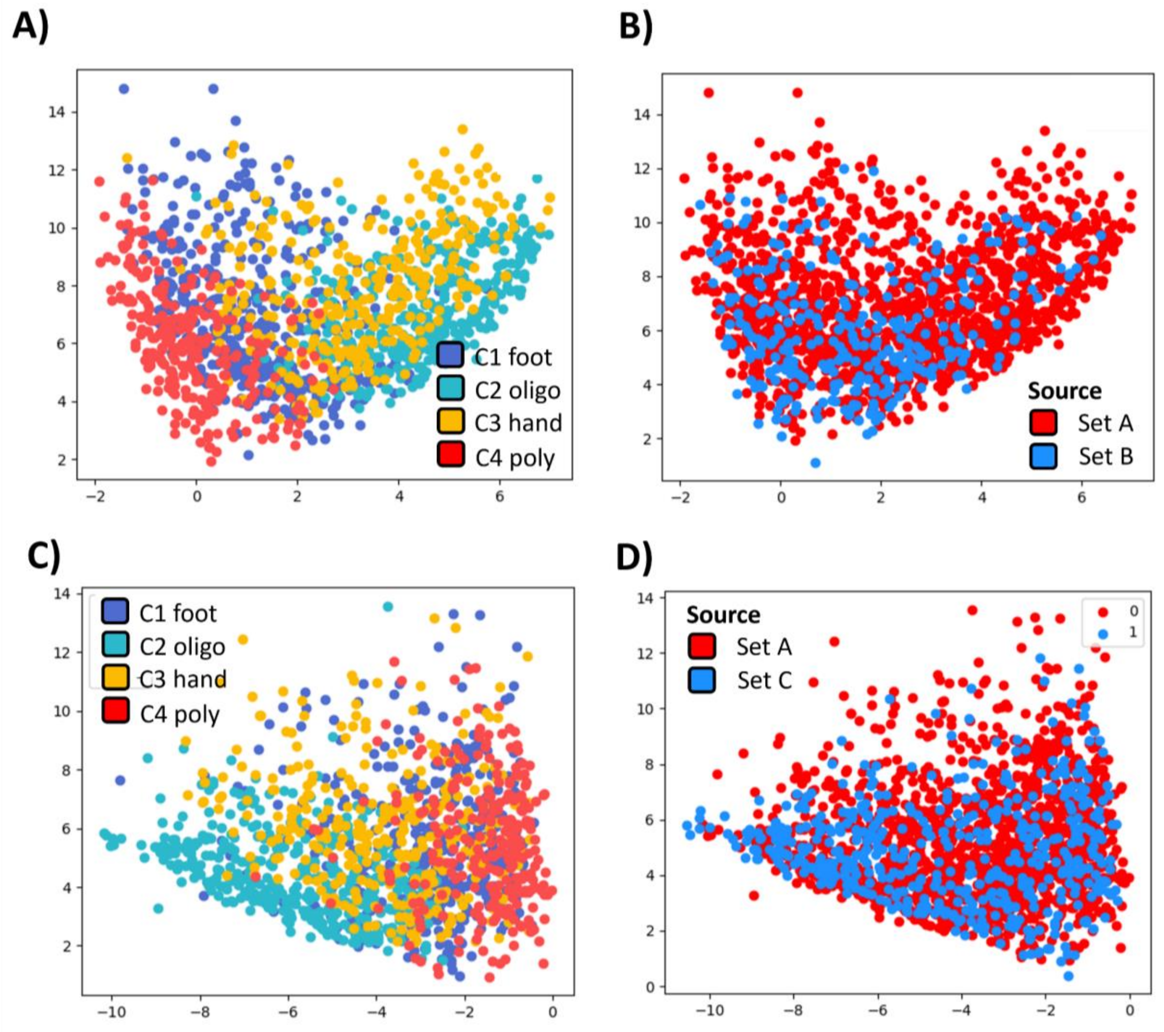


***Supplementary Fig. S27: The 2D UMAP representation of multimodal autoencoder (MMAE), where each point represents a patient.*** *Here, we see the distribution of clusters across the shared product space with Set B (A), and in complement the orientation of the newly projected patients in blue vs original patients in red (B). Similarly, we report the shared product space for set C (C), and the orientation of the new patients in this patient embedding (D).*

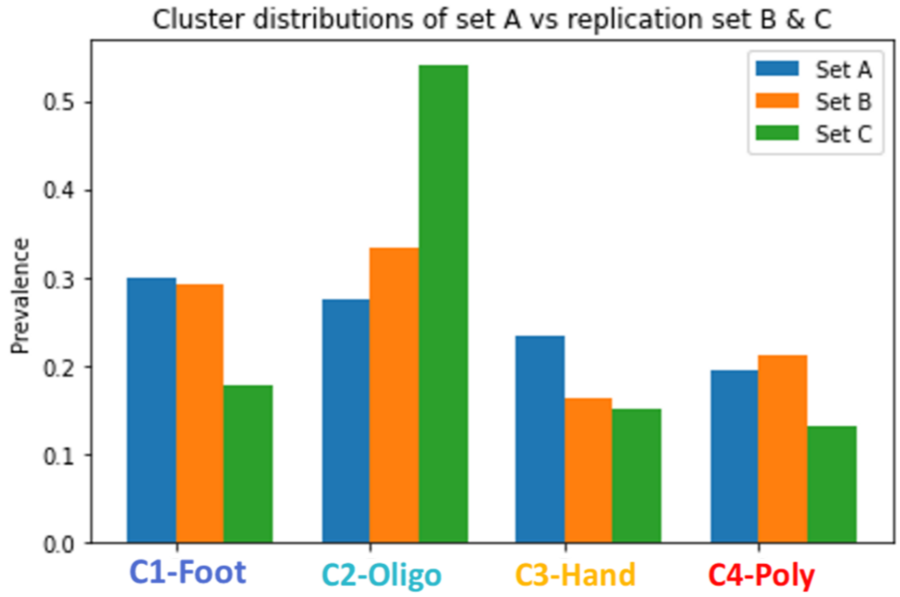


*Supplementary Fig. S28: Spatial variance of patients compared between the development (set A) and replication set (set B and C).*

### A) Methotrexate Failure

| | *Power analysis* |
|---|---|
| *Cluster prevalence (C3)* | 0.53 |
| *Cluster prevalence (C4)* | 0.47 |
| *Probability exp. group (C3)* | 0.837 |
| *Probability control group (C4)* | 0.697 |
| *Postulated HR* | 0.48 |
| *alpha* | 0.05 |
| *Technique* | Sample size calculation under CPH |

| | *N* | *MTX-starter* |
|---|---|---|
| *C1-Foot* | 91 | 69 |
| *C2-Oligo* | 279 | 214 |
| *C3-Hand* | 77 | **59** |
| *C4-Poly* | 68 | **51** |

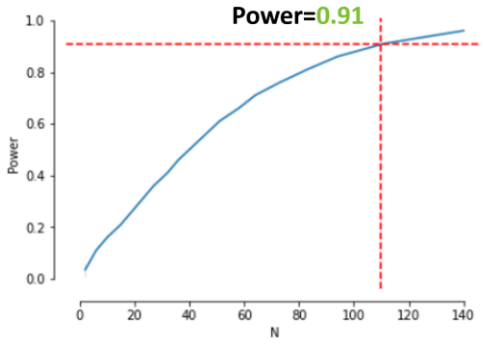


### B) Remission

| | *Power analysis* |
|---|---|
| *Cluster prevalence (C3)* | 0.48 |
| *Cluster prevalence (C4)* | 0.52 |
| *Probability exp. group (C3)* | 0.48 |
| *Probability control group (C4)* | 0.66 |
| *Postulated HR* | 1.76 |
| *alpha* | 0.05 |
| *Technique* | Sample Size calculation under CPH |

| | *N* | *Remission info* |
|---|---|---|
| *C1-Foot* | 91 | 46 |
| *C2-Oligo* | 279 | 186 |
| *C3-Hand* | 77 | **30** |
| *C4-Poly* | 68 | **32** |

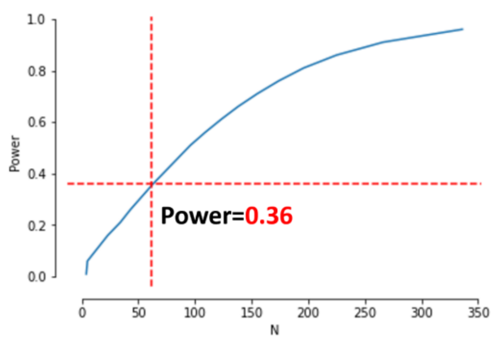


*Supplementary Fig. S29: Power curves for Reumazorg Zuid West Nederland hospital data based on postulated hazard ratio between C3 and C4 for Methotrexate-failure.*
